# COMBINING CHEST X-RAY AND CLINICAL COVARIATES FOR OSTEOPOROSIS SCREENING: A MULTIMODAL DEEP LEARNING APPROACH

**DOI:** 10.64898/2026.09.03.26362168

**Authors:** Vladimer Kobayashi, James Punsalan, Gabriel Teodoro Castellano Baluyut

## Abstract

Osteoporosis is a major global health concern, yet screening remains inaccessible in many economically challenged and low-resource settings due to the high cost and operational complexity the gold standard Dual-energy X-ray Absorptiometry (DXA) scan requires. This study presents a multimodal deep-learning approach for opportunistic osteoporosis screening using chest X-rays and clinical variables such as age and gender. A total of 406 samples were retrospectively collected from a national hospital, comprising annotated chest radiographs paired with their DXA results from patients 50 and above. Two convolutional neural network (CNN) architectures, ResNet50 and EfficientNetb3, were deployed under three configurations: image-only, early fusion, and late fusion. Image preprocessing includes CLAHE+Gamma Correction, followed by resizing, local regions of interest (ROI) extraction, and augmentation. The multimodal integration was conducted via early fusion (pixel-level) and late fusion (feature-level), with clinical variables normalized and encoded into a 1×3 vector. A custom Residual MLP classifier was employed in the late fusion models. Hyperparameter tuning was performed using Optuna’s Bayesian optimization. Evaluation metrics included Accuracy, AUC, Precision, Recall, Sensitivity, and F1 Score. Among all models, Late Fusion EfficientNetb3 achieved the best performance on the global chest X-ray, with an AUC of 72.73%. Moreover, EfficientNetb3 consistently outperformed the ResNet50 architecture across all local ROI, Right Clavicle, Right Scapula, and Spine, with fusion models yielding higher sensitivity and balanced classification. The framework achieved competitive diagnostic performance despite the limitations of chest radiographs and an imbalanced dataset. Moreover, the findings demonstrated the feasibility of integrating chest X-rays with clinical covariates for AI-assisted osteoporosis screening, aiming to fill the accessibility gaps in preventive bone health diagnostics.

## 1 INTRODUCTION

### 1.1 Background of the Study

Osteoporosis is a skeletal disease characterized by decreasing density of the bones due to the inadequate calcium content (Jang et al., 2022). This ‘silent epidemic’ has inflicted over 200 million individuals worldwide with the Philippines having a prevalence rate of 11.3% in females and 9.0% in males (Li-Yu, 2007; Wang et al., 2021). As the population ages and life expectancy increases, the number of individuals at risk for osteoporosis continues to grow, particularly among those aged 60 and above.

Osteoporosis is diagnosed using Dual-energy X-ray Absorptiometry (DXA) scan, with the World Health Organization (WHO) classifying T-scores of –2.5 or lower as indicative of osteoporosis, and T-scores between –1.0 and –2.5 as indicative of osteopenia which is a condition of reduced bone density that may progress to osteoporosis (Ho et al., 2021). While DXA scan is the current ‘gold standard’ for diagnosing osteoporosis, this procedure can be inaccessible in some low resource settings because of its high cost and sophisticated operational demands (Ashames et al., 2021).

The integration of Artificial Intelligence (AI) in patient care has the potential to ease the diagnostic process through trained AI models, specifically in Neuroradiology, Oncological Imaging, Cardiovascular Imaging, and Abdominal Imaging, highlighting that AI can be a vital piece in improving diagnostic precision (Najjar 2023).With its promised potential, the medical imaging field is seeing a shift toward AI-assisted diagnostics. Studies have shown that several characteristics seen in routine medical imaging, including alterations in bone structure or vertebral fractures, might function as visual indicators of osteoporosis (Ho et al., 2021; Wang et al., 2021; Yamamoto et al., 2020). Arguably, X-ray emerges as a viable method for screening osteoporosis, offering a complementary approach to standard DXA scans. Studies have specifically focused on using radiographic data to screen for osteoporosis, examining areas such as the hip (Yamamoto et al., 2020), foot (Ashames et al., 2021), spine (Lee et al., 2020), and chests (Jang et al., 2022; Wang et al., 2021).

Furthermore, the cost-effectiveness of X-ray scans compared to DXA scans strengthens the case for their use as a screening tool. The Lung Center of the Philippines (2024) lists the cost of DXA scans two times higher compared to the cost of standard Chest X-ray scans (which is typically twice the minimum daily wage). Additionally, X-ray scans require less specialized equipment and training compared to DXA. As X-ray scans are considered a routine procedure in many medical examinations, they do not incur any additional cost for the patient and can be utilized as an opportunistic screening procedure for osteoporosis.

A number of studies have demonstrated the potential of X-ray for osteoporosis screening. For example, a study utilized a multi-regions of interest (ROI) of local and global bone structure from the chest X-ray to predict bone mineral density (BMD) and diagnose osteoporosis (Wang et al. 2021). Another paper proposed a screening model called OsPor-screen that uses supervised deep learning (DL) to identify osteoporosis (M. Jang et al. 2022).

Other researchers have introduced a multimodal approach via late fusion to integrate radiographic and patient clinical data in their screening process (Sukegawa et al., 2022 ; Yamamoto et al., 2020; Yamamoto et al., 2021). They demonstrated an incremental improvement of AUC scores when integrating clinical covariates into the image-only model with improvements ranging from 2.1% to 3.2% across the said studies. Age, gender, and body mass index (BMI), have been identified as clinically important risk factors in osteoporosis diagnosis (Sukegawa et al., 2022; Yamamoto et al., 2020), the improved performance metrics on the said studies suggest that adding clinical variables can incrementally improve osteoporosis screening.

While these advancements have been happening globally, there is a notable gap in published studies utilizing Southeast Asian demographics, specifically in the Philippines on the diagnosis of osteoporosis through chest X-ray radiographs and clinical data. This underscores the need for tailored models that can account for the demographic and health profiles in the Philippines (Pouresmaeili et al., 2018). With this study, we can ensure the developed models are optimized for local healthcare practices and population-specific characteristics, which may differ significantly from those models developed in other countries. This is crucial because models built on non-local data can lead to less accurate or less effective healthcare interventions when applied to different populations (Futoma et al., 2020; Pouresmaeili et al., 2018). These presented gaps highlight the opportunity to leverage current advancements to address local healthcare challenges effectively.

This study explored the use multimodal deep learning approach that combines chest X-ray radiographs and key clinical variables such as age, gender, and BMI to screen for osteoporosis. CNN was used to handle the X-ray radiographs as it is capable of handling the complexities of pixel-based data, making it particularly adept at automatically learning spatial hierarchies of features directly from images. This capability of CNN makes them suitable for medical imaging tasks where diagnoses often rely on subtle visual cues on an image. Moreover, with the integration of clinical variables in the CNN, we utilized and compared two multimodal approaches, namely, early and late fusion (Pei et al., 2023). For this study, firstly the appropriate CNN architecture was selected and secondly different strategies on how to integrate the clinical covariates were compared. By testing and comparing various models, the most efficient and most accurate approach for osteoporosis screening can be identified. Given these considerations, we sought to answer the following research questions:

1. Which Convolutional Neural Network (CNN) architecture yielded the best performance for osteoporosis screening using chest X-ray images?
2. Does the integration of clinical variables such as age, and gender improve the performance of image-only CNN models?
3. Which fusion strategy (early or late) yields better performance in a multimodal deep learning approach to osteoporosis screening?

### 1.2 Objective of the Study

Based on the preceding research questions the following objectives were formulated: (1) apply and compare two CNN architectures for radiographic data; (2) apply and compare two multimodal approaches in integrating both radiographic and clinical data; and (3) compare the performance of the standalone CNN architecture against those integrated within the ensemble model.

### 1.3 Benefits and Contributions

This study aligns with the United Nations Sustainable Development Goals, particularly SGD 3: Good Health and Well-being, and SGD 9: Industry, Innovation, and Infrastructure. By improving screening accessibility, this study contributes to better health outcomes and well-being for individuals at risk. Moreover, introducing this innovative take on medical diagnostics highlights the potential of technology in enhancing the healthcare infrastructure and fostering innovation within the industry, contributing to building a sustainable and resilient healthcare system.

Economically, the implementation of this multimodal approach could decrease the financial burden on healthcare systems as it answers the need for more accessible screening methods. In terms of methodological contributions, the study contributes to the field by demonstrating the efficacy of integrating machine-learning techniques with radiographic and clinical data. This approach can serve as a blueprint for other researchers aiming to capitalize on artificial intelligence in healthcare, particularly in developing countries where healthcare resources and data could be scarce.

## 2 REVIEW OF RELATED LITERATURE

### 2.1 Artificial Intelligence (AI) in Medical Imaging

AI, with its recent resurgence, has been a crucial technological development with the potential in medical diagnostics. It is instrumental in interpreting medical images such as X-ray radiography, Computed Tomography (CT) scans, and Magnetic Resonance Imaging (MRI) scans, aiding in detecting and diagnosis of diseases such as breast cancer, lung nodules, brain tumors, fractures, and cardiovascular diseases (Lee et al., 2020).

While manual review and experience-based decisions by medical professionals have been the traditional diagnostic methods for years, the integration of AI and ML through convolutional neural networks (CNNs) is becoming a new standard of care (Najjar, 2023; Soffer et al., 2019). CNN, a class of deep learning (DL) models that underlies many image recognition and segmentation models, is capable of learning complex patterns, enabling high accuracy scores in visual tasks across various radiological applications.

The impact this technological advancement had in the field of medicine have been examined in a series of studies documenting that the integration of AI and ML models was of great help in patient care (Najjar 2023). For instance, integrating AI and ML models into various aspects of cancer imaging could streamline cancer care, including risk assessment, screening, diagnosis, and response evaluation (Koh et al., 2022). Moreover, in cardiovascular and abdominal imaging, AI-driven models introduced a new standard in disease detection and monitoring (Zhou et al., (2021). Another study accounted for success in applying AI models to detect and analyze gastrointestinal and hepatic diseases (Kumar et al. 2021)

### 2.1.1 Convolutional Neural Network for Radiograph Evaluation

A study which explored the possibility of predicting Bone Mineral Density (BMD) relative to Dual-energy x-ray Absorptiometry (DXA) from patient radiographs, has introduced a novel approach that utilized a modified CNN model, named ‘DeepDXA’ (Ho et al., 2021). DeepDXA was developed to perform classification tasks on identifying patients with osteopenia and osteoporosis based on T-score thresholds. The model yielded a sensitivity of 94% and a specificity of 65% for detecting osteopenia, and a sensitivity of 76% and a specificity of 87% for osteoporosis, showing its efficacy in identifying these conditions. A similar study explored the potential of Deep Learning Neural Networks (DLNN) models to predict BMD from unenhanced abdominal CT images, specifically targeting lumbar vertebrae (Yasaka et al. 2020). The researchers have utilized a different imaging modality, exploring CT scans as viable imaging sources for BMD assessment. Their study utilized a supervised training of CNN with axial CT images of lumbar vertebrae (L1–4), contrasting with Ho et al.’s approach that employed anteroposterior pelvic X-rays.

The results demonstrated a significant correlation between the estimated BMD values from the CNN model and the actual BMD values from DXA. It had Pearson correlation coefficients of 85.2% and 84.0% for internal and external validation datasets, respectively. Moreover, the ability to diagnose osteoporosis using the CNN-derived BMD values (BMDCNN) was encouragingly high, with area under the curve (AUC) scores of 96.5% and 97.0% for the internal and external datasets, respectively. These findings showcases deep learning model’s efficacy in osteoporosis detection directly from CT images.

In osteoporosis screening AI and ML models were employed to predict Bone Mass Density (BMD) using simple spine X-rays, using a combination of VGGnet and classification by random forest based on the maximum balanced classification rate (BCR) for classification (Lee et al. 2020). Digging deeper into radiology, a growing interest in researching the utilization of CNN as an aid in interpreting X-ray images has been seen for the past years. This section dissects these different approaches, compare and contrast them to determine which approach is a potential candidate model to be deployed in this study.

Moreover, more recent studies explored the use of CNN Models to assess chest (Jang et al., 2022; Wang et al., 2021) and hip (Yamamoto et al., 2020) X-ray radiographs. Here, they utilized various CNN architectures such as RestNet, GoogleNet, EfficientNet, AlexNet, MobileNetV2, and InceptionV3 architectures that showed improved prediction accuracy and efficiency (Ho et al., 2021; Yamamoto et al., 2020, 2021).

### 2.1.2 AI-assisted diagnosis of Osteoporosis using Chest X-rays

As demonstrated in previous studies, osteoporosis can be potentially screened through typical X-ray radiographs, hence, there is a growing interest in AI-assisted diagnosis specifically using Chest X-rays (Jang et al., 2022; Wang et al., 2021).

A deep learning model, OsPor-screen can determine the presence of Osteoporosis with a reported AUC of 88% in external tests which included the gradient-weighted class activation mapping (Grad-CAM) for visualizing and interpretating the model’s predictions (M. Jang et al. 2022). A more complex method involved automatically extracting multiple specific regions of interest within the available Chest X-ray which are then fed to a CNN Architecture, VGG-16, that yielded a correlation coefficient of 84.0% and an AUC of 93.6% in comparison to the DXA measurements (Wang et al. 2021).

A paper reviewed 40 studies via a two-step literature search using the PubMed and Web of Science databases (He et al. 2024). The review included studies focused on routine radiologic methods such as X-ray, CT scans and Magnetic Resonance Imaging (MRI) used to screen for osteoporosis. Notably, the paper highlighted that integrating clinical covariates into the model improves AUC scores by 2%-4%. These observations were supported by another review study which reported high diagnostic accuracy (Yen et al.,2024). Furthermore, the use of pre-trained models, data augmentation, and image standardization enabled studies with limited datasets to achieve good performance metrics.

Despite promising results, they underscored a need for additional prospective multicenter studies involving diverse patient populations to confirm the acceptability of these techniques in the clinical setting.

Another study that demonstrated the potential of AI in Analyzing Radiology was focused on validating a developed deep learning model, FORM, in predicting hip fracture risk from both X-ray and CT scan images (Schmarje et al., 2022). The results highlighted the effectiveness of FORM, outperforming conventional methods like the Cox Proportional-Hazards Model and FRAX. Furthermore, the integration of patient demographics and clinical history with image data were deemed as contributing factors in improving the performance of the model.

These studies, encompassing a range of methodologies from deep learning models such as DeepDXA and various image enhancement techniques, have shown the impact of AI in improving diagnostic accuracy for conditions such as osteoporosis and COVID-19.

### 2.2 Multimodal approach: Integrating Key Clinical Variables

Research has underscored the role of integrating multiple data modalities in enhancing diagnostic accuracy and treatment efficacy (Pei et al. 2023). They highlighted three key fusion approaches employed in multimodal learning: early fusion, late fusion, and hybrid fusion.

*Early fusion* involves combining different data types at the input stage before the training process. *Late fusion* merges these different data types at a later stage, typically at the decision level. *Hybrid fusion* offers a more flexible approach as it allows some of the modalities to be integrated in the early stage while reserving the others for later stages. Shown in Figure 1 is the schematic diagram of the three multimodal approaches.

**Figure 1.**
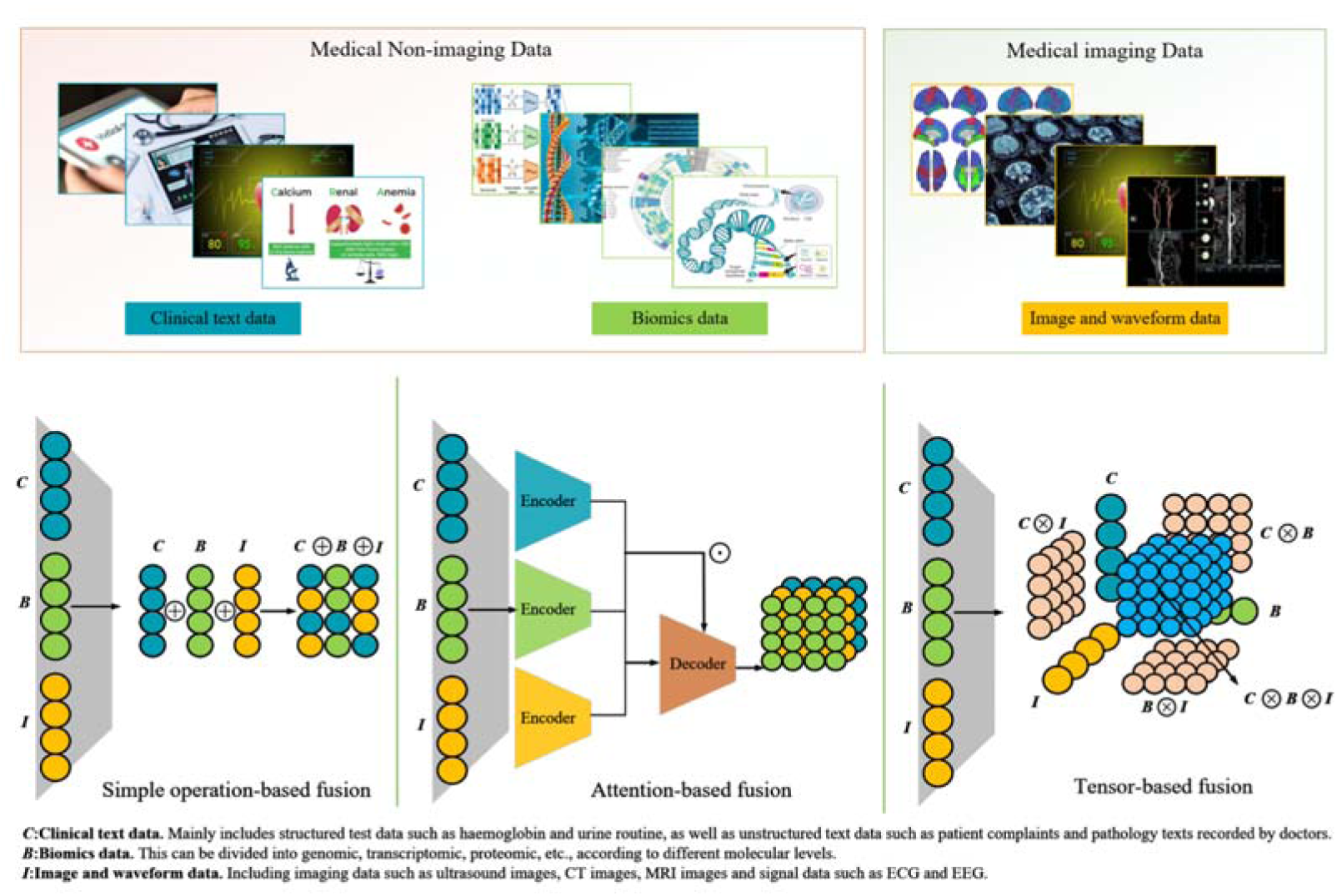
Schematic Diagram of Early Fusion (Simple operation-based), Late Fusion (Attention-based) and Hybrid Fusion (Tensor-based) (Lifted from Pei et al. (2023).

The integration of key clinical variables for chest X-ray-based diagnosis is rarely explored (Yamamoto et al. 2020; Yamamoto et al. 2021; Sukegawa et al. 2022).

The ensemble deep-learning models developed by a number of researchers included Age, BMI, and gender (Yamamoto et al. 2020; Yamamoto et al. 2021; Sukegawa et al. 2022). The processed image data from the CNN, along with the merged clinical variable data, were fed into the fully connected layer. The final model for identifying osteoporosis made its predictions using the rectified linear unit (ReLU) activation function. integrating key clinical variables led to improved diagnostic accuracy.

Table 2 provides a comparative overview of the AUC scores achieved by ResNet50 and EfficientNetB3 across various studies that integrated clinical data with medical images through ensemble models. These prior studies demonstrated that incorporating clinical covariates led to noticeable improvements in model performance, yielding effect sizes ranging from 2.1% to 3.2%. This emphasizes the tangible value of multimodal learning in enhancing diagnostic accuracy, particularly in osteoporosis screening where radiographic indicators may not always be visually distinct.

**Table 1.** Summary of studies that utilized radiographic images in screening osteoporosis (Lifted from He et al. (2024)).

| Author | Country | Year | Objective | Material | Dataset | Algorithm |
| --- | --- | --- | --- | --- | --- | --- |
| Breit et al. | Switzerland | 2023 | BMD prediction | CT | 109 patients | DI2IN |
| Chen et al. | Taiwan (China) | 2023 | Osteoporosis diagnosis | CT | 197 patients (External validation: 397) | ResNet-50, SVM |
| Derkatch et al. | Canada | 2019 | Fracture risk prediction | DXA | 12742 patients | InceptionResNetV2, DenseNet |
| Dzierzak et al. | Poland | 2022 | Osteoporosis diagnosis | CT | 100 patients (Training: Validation: Test = 2:1:1) | VGG-16, VGG-19, MobileNetV2, Xception, ResNet-50, InceptionResNetV2 |
| Fang et al. | China | 2020 | Osteoporosis diagnosis | CT | 1449 patients (Training: 586 patients, Test: 863 patients) | DenseNet-121, U-net |
| Ho et al. | Taiwan (China) | 2021 | BMD prediction | X-ray | 3472 images | ResNet-18 |
| Hong et al. | South Korea | 2023 | Osteoporosis diagnosis | X-ray | 9276 patients | EfficientNet-B4 |
| Jang et al. | South Korea | 2022 | Osteoporosis diagnosis | X-ray | 1089 chest radiographs (Training:Validation:Test = 7:1:2) | CNN |
| Jang et al. | South Korea | 2022 | Osteoporosis diagnosis | X-ray | Total: 1001 patients (Training:Validation:Test = 8:1:1, External validation: 117 patients) | NLNN |
| Kong et al. | South Korea | 2022 | Fracture risk prediction | X-ray | 1595 participants (Training: 1416 participants, Test: 179 participants) | DeepSurv |
| Lee et al. | South Korea | 2018 | Osteoporosis diagnosis | DPR | 1268 patients (Training and validation: 1068, Test: 200 patients) | MC-DCNN, SC-DCNN |
| Lee et al. | South Korea | 2020 | Osteoporosis diagnosis | DPR | 680 patients (Test dataset: 20%) | CNN-3, VGG16, VGG-16_TF, VGG-16_TF_FT |
| Lee JS et al. | UK | 2019 | Fracture detection | Panoramic radiography | Total: 1001 participants | CNN |
| Liu et al. | China | 2019 | Osteoporosis diagnosis | X-ray | 89 patients | BP network, SVM, U-net |
| Löffler et al. | Germany | 2021 | Osteoporosis diagnosis | CT | 192 patients | CNN |
| Mao et al. | China | 2022 | Osteoporosis diagnosis | X-ray | 5652 patients (Training:Validation:Test set 1:Test set 2 = 8:1:1:1) | DenseNet |
| Monchka et al. | Canada | 2021 | Fracture risk prediction | DXA | 12742 patients | InceptionResNetV2, DenseNet |
| Nguyen et al. | South Korea | 2021 | BMD prediction | X-ray | 330 patients (660 hip X-ray images) (Training: 510 images, Test dataset: 150 images) | VGGNet |
| Pan et al. | China | 2020 | BMD prediction | CT | 374 patients | U-net |
| Pickhardt et al. | USA | 2022 | Osteoporosis diagnosis | CT | 11035 patients | TernausNet |
| Rühling et al. | Germany | 2021 | BMD prediction | CT | 193 patients (Training:Test = 8:2) | 2D DenseNet, 3D DenseNet |
| Sato et al. | Japan | 2022 | BMD prediction | X-ray | 10102 patients (Training:Validation:Test = 7:2:1) | ResNet-50 |
| Sollmann et al. | Germany | 2022 | BMD prediction | CT | 144 patients | CNN |
| Sukegawa et al. | Japan | 2022 | Osteoporosis diagnosis | DPR | 778 patients | EfficientNet-b0, b3, b7, ResNet-18, 50, 152 |
| Tang et al. | China | 2020 | BMD prediction | CT | 213 patients (Training: 150 patients, Test: 63 patients) | Net, DenseNe |
| Tariq et al. | USA | 2022 | Osteoporosis diagnosis | CT | 6083 images (Prospective test group: 344 patients) | DenseNet-121 RF |
| Tomita et al. | USA | 2018 | Fracture detection | CT | 1432 cases<br>(Training: 1168 cases,<br>Validation: 135 cases,<br>Adjudicated test set: 129 cases) | ResNet-34, LSTM |
| Uemura et al. | Japan | 2022 | BMD prediction | CT | 75 patients | U-net |
| Wani et al. | India | 2022 | Osteoporosis diagnosis | X-ray | 240 patients | ResNet-18, AlexNet, VGG-16, VGG-19 |
| Xiao et al. | USA | 2022 | Fracture detection | Chest X-ray | 6674 cases<br>(Training: 5970 cases, Test: 704 cases) | Not mentioned |
| Yamamoto et al. | Japan | 2020 | Osteoporosis classification | X-ray | 1223 patients | ResNet-18, ResNet-34, GoogleNet, EfficientNet-b3, EfficientNet-b4 |
| Yamamoto et al. | Japan | 2021 | Osteoporosis classification | X-ray | 1699 patients | ResNet-18, 34, 50, 101, and 152 |
| Yasaka et al. | Japan | 2020 | BMD prediction | CT | 183 patients | CNN |
| Zhang et al. | China | 2023 | Osteoporosis classification | CT | 1048 patients<br>(Training: Validation: Test = 5:1:4) | U-net |
| Zhang et al. | China | 2020 | Osteoporosis diagnosis | X-ray | Training and internal validation: 910 patients, Test dataset 1: 198 patients, Test dataset 2: 147 patients | DCNN |
| Zhao et al. | China | 2022 | Osteoporosis diagnosis | MRI | Training: 142 patients, Validation: 64 patients, External validation: 25 patients | U-Net, LASSO |

**Table 2.** Summary of AUC scores of ResNet50 and EfficientNetb3 on image-only and ensemble models (Lifted from Sukegawa et al. (2022) and Yamamoto et al. (2020, 2021)).

| CNN Architecture | Author | Medical Images | AUC Score |  | Effect Size |
| --- | --- | --- | --- | --- | --- |
|  |  |  | Image-only | Ensemble |  |
| ResNet50 | Yamamoto et al. (2021) | Hip X-ray | 88.5% | 90.6% | 2.1% |
| EfficientNetb3 | Yamamoto et al. (2020) | Hip X-ray | 90.9% | 93.7% | 2.8% |
|  | Sukegawa et al., 2022) | Panoramic X-ray | 86.7% | 89.9% | 3.2% |

While the benefits of multimodal fusion are evident, existing literature primarily explored this approach using imaging data from the hip or panoramic dental radiographs. There remains a significant gap in leveraging chest X-rays, a more widely available and routinely performed imaging modality, for multimodal osteoporosis screening. This study aimed to address this gap by evaluating early and late fusion strategies on chest X-rays combined with age and gender data, thereby contributing to the growing interest in localized, accessible, and effective AI-assisted diagnostic tools for osteoporosis.

## 3 MATERIALS AND METHODS

This section presented the research design for developing a multimodal deep learning-based model to screen for osteoporosis by integrating chest X-rays with key clinical variables, namely age and gender. The processes of data collection, preprocessing, model training, and evaluation were outlined and illustrated in Figure 2.

**Figure 2.**
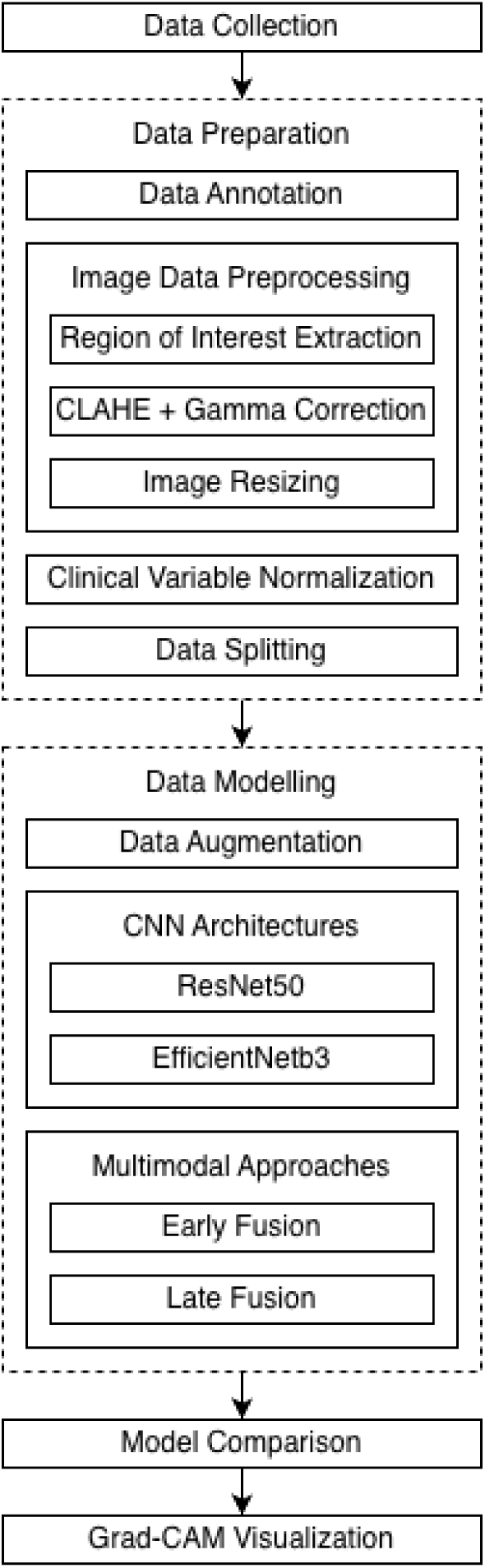
Methodological Framework of the Study.

### 3.1 Data Collection

Archival Chest X-ray and DXA data from patients aged 50 years old and above between January 2014 and January 2022 were retrospectively collected from health records at The Medical City Hospital in Pasig City, Philippines. The study included only those patients whose chest X-ray images were in anteroposterior (AP) or posteroanterior (PA) views, and who had DXA results done within six months from taking the X-ray images. Moreover, the study excluded patients with visible non-biologic hardware or devices such as endotracheal tubes, pacemakers, or sternal wires, along with patients with chest X-rays done in apicolordotic or lateral views only, and patients whose chest X-ray and DXA scan were done beyond the six months apart. Ethical clearance was secured through The Medical City’s Institutional Review Board.

### 3.2 Data Preparation

To ensure that the data input into the multimodal deep learning model for the diagnoses of Osteoporosis are consistent, several data preparation techniques before the classification was employed. The following preprocessing techniques were done for the chest Xray images: image region of interest extraction, CLAHE + gamma correction, image resizing were implemented to standardize the input data. Moreover, clinical data variables were normalized and encoded. For model training we split the dataset as follows: eighty percent (80%) were used for the training, ten percent (10%) were for validation, and the other ten percent (10%) were reserved for testing the model (Sukegawa et al. (2022) ; Yamamoto et al.’s 2021). X-rays were labelled by a radiologist from The Medical City hospital. Each X-ray was categorized as either Normal, Osteopenia, or Osteoporosis, by pairing it with the patient’s DXA results following the WHO criterion. The DXA results served as the reference standard, or ground truth, for training the Deep Learning Model.

### 3.3 Image Data Preprocessing

Prior to model training, the collected Chest X-rays underwent a series of preprocessing.

### 3.4 Image Region of Interest Extraction

We extracted the Right Clavicle, Right Scapula, and Spine as our local ROIs. Among the evaluated regions, these three anatomical sites were selected based on their relevance and potential discriminative performance (Wang et al.’s 2021). For completeness, the global ROI image, which is the entire chest X-ray image, was also analyzed. Figure 3 shows the parts of the chest X-ray that were identified as the ROIs.

**Figure 3.**
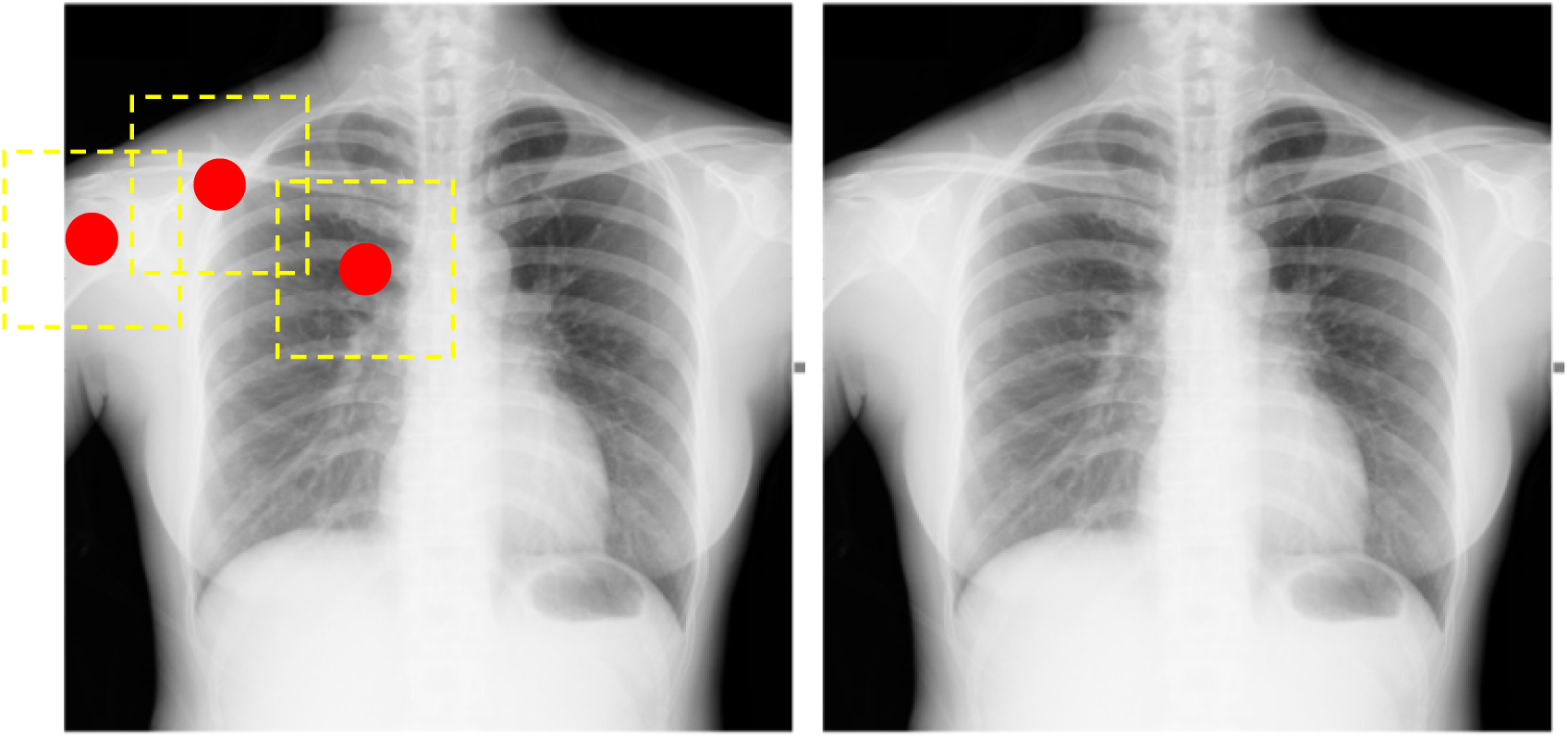
The identified ROIs include the Right Clavicle, Right Scapula, and Spine as the Local ROI (left image), and the entire Chest as the Global ROI (right image) (Lifted from Wang et al.’s (2021)).

To execute this, we deployed a predefined algorithm that uses the PSPNet model (Cohen et al. 2021), a model renowned for its efficacy in semantic segmentation tasks, which outputted a set of pixel-wise classification maps. These maps indicated the likelihood of each pixel belonging to a specific anatomical structure.

A sigmoid function was then applied to yield a probability map, which served as the threshold for the binary masks that isolated the Right Clavicle, Right Scapula, and Spine areas from the rest of the chest anatomy. After masking, the 512 × 512-pixel image was exported back to the repository, where it underwent the rest of the preprocessing process.

### 3.5 Image CLAHE + Gamma Correction

Another issue lies in the inherent visual challenges of interpreting standard chest X-rays for osteoporosis screening. Differentiating osteopenic cases from normal ones can be difficult due to overlapping anatomical structures, low contrast, and weak bone boundary representation, which may hinder the model’s ability to extract relevant features for classification (Huang et al., 2025).

Since classification performance is often affected by image quality, we opted to use the Contrast-Limited Adaptive Histogram Equalization (CLAHE) applied in conjunction with Gamma Correction (GC) to improve local contrast and overall brightness levels of the chest radiographs best-performing (Rahman et al.’s 2021).

CLAHE was used to highlight fine anatomical details, such as trabecular bone patterns, cortical bone thickness, and vertebral endplates, which are critical for osteoporosis screening. After CLAHE, Gamma Correction was applied as a nonlinear enhancement technique, adjusting pixel luminance through a gamma mapping function to emphasize important bone features. Gamma values ranging from 0.5 to 3.5 were tested to assess their impact on the visibility of anatomical structures.

The resulting gamma-adjusted pixel value,*g(x)*, can be represented as

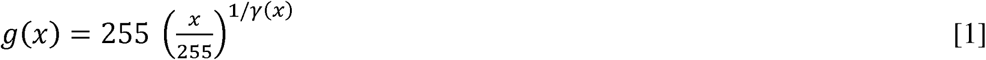

where 1/*y(x)*is a dynamic function derived from projection mappings defined in the original study.

### 3.6 Image Resizing

After gamma correcting the extracted images, they were then resized to comply with the dimensional requirements of the CNN models, that is, 224 × 224 pixels for ResNet50 and 300 × 300 pixels for EfficientNetB3. Given that the images were in greyscale mode, we used bilinear interpolation to preserve their quality. This method balances computational efficiency and image quality by preserving details and reducing visual artifacts, making it suitable for deep learning tasks (Rukundo 2023). Moreover, this method works by interpolating pixel values by using linear interpolation first in one direction, and then in the other, as represented by Figure 4.

**Figure 4.**
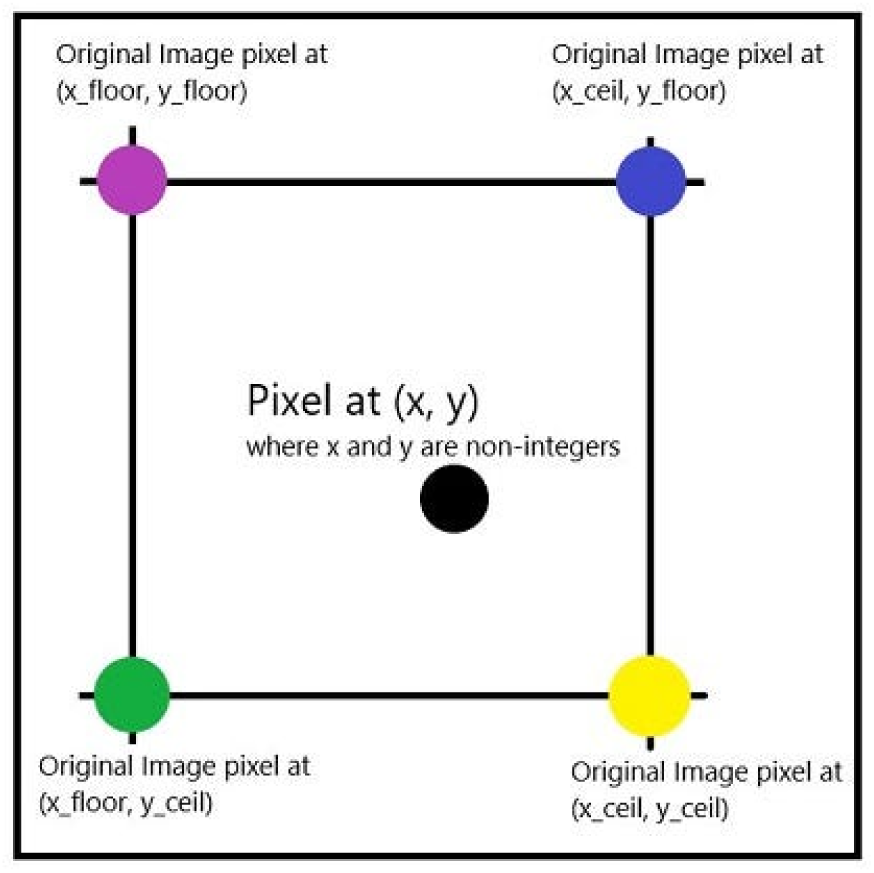
Pixels from the original image surrounding pixel at (x,y) (Lifted from Darji (2021)).

Where we let,

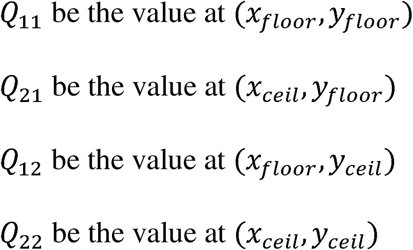

To interpolate value at *Q* at (x, y) we perform the following steps:

1. Horizontal interpolation

1.1. Interpolate *between Q_11_and Q_21_to* find an intermediate value

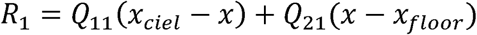

1.2. Interpolate between *Q_12_*and *Q_22_* to find an intermediate value

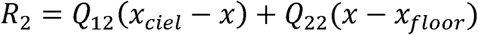

2. Vertical Interpolation

2.1. Interpolate between *R_1_* and *R_2_*using the y coordinates to find the final value

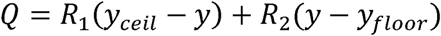

Following resizing, the images underwent normalization to scale the pixel values to a standard range that will aid in the convergence of the network during the training phase. The Z-score Normalization technique is represented by

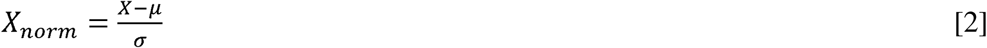

where,

*X*: original pixel value

**μ**: mean of the pixel values

σ: standard deviation of the pixel values

#### Clinical Variable Normalization

For the clinical variables, we applied the following preprocessing techniques (Yamamoto et al. 2021).

#### Age Normalization

Mean normalization was deployed to ensure that these features are on a similar scale as the CNN-derived features. This method adjusts each value by subtracting the mean of the dataset and then dividing by the standard deviation as represented in equation [3]

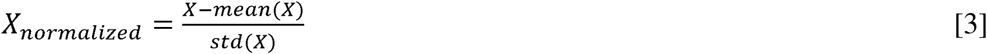

Where,

*X* is the original value.

#### Gender Encoding

The gender variable was encoded using the one-hot vector representation. Given that gender variables only include male and female, the process converts the variable into binary vector format, where male is represented as [1,0], and female is represented as [0,1].

#### Combining into a 1 × 3-dimensional vector

We combined the normalized age, BMI, and encoded gender into a 1 × 3-dimensional vector.

#### Data Modelling

##### Data Augmentation

Data augmentation can enhance the performance of deep learning models by increasing data diversity, reducing overfitting, and improving generalization, especially in cases where gathering new data is costly or not feasible (Mikołajczyk-Bareła & Grochowski, 2018; Rahman et al., 2021). Since we have a relatively small dataset, we deployed data augmentation to produce artificial data that could simulate plausible variations in real life. We adopted the following data augmentation techniques (Ho et al.’s 2021):

##### Rotation

Each image was randomly rotated by an angle between -20° and +20°.

##### Scaling

Each image was randomly scaled with a factor ranging from 0.8 to 1.2.

##### Shearing

This was applied with angles between -15° and +15°.

##### Translation

Each image was translated both vertically and horizontally by up to 10% of the image size.

##### Horizontal Flipping

Each image has a 50% of being flipped horizontally.

Deploying these five data augmentation techniques increased the number of training data, as shown in Table 3.

**Table 3.**
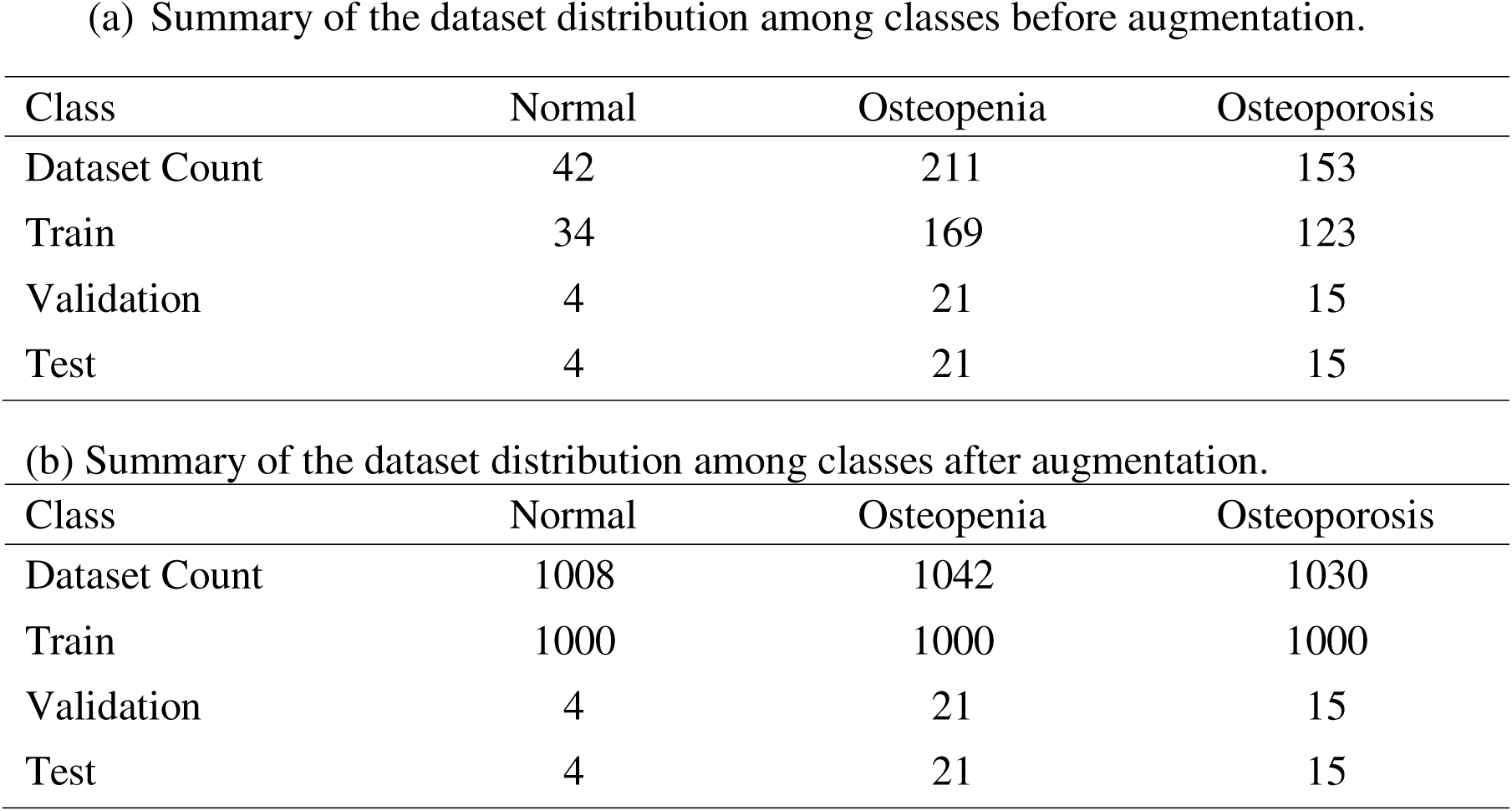
Training, validation and test datasets: (a) before data augmentation and (b) after data augmentation.

(a) Summary of the dataset distribution among classes before augmentation.
| Class | Normal | Osteopenia | Osteoporosis |
| --- | --- | --- | --- |
| Dataset Count | 42 | 211 | 153 |
| Train | 34 | 169 | 123 |
| Validation | 4 | 21 | 15 |
| Test | 4 | 21 | 15 |

(b) Summary of the dataset distribution among classes after augmentation.
| Class | Normal | Osteopenia | Osteoporosis |
| --- | --- | --- | --- |
| Dataset Count | 1008 | 1042 | 1030 |
| Train | 1000 | 1000 | 1000 |
| Validation | 4 | 21 | 15 |
| Test | 4 | 21 | 15 |

#### CNN Architectures

For the training of our models, we utilized a selection of pre-trained CNN architectures, each chosen for its specific strengths and performance in medical imaging tasks (Ho et al., 2021; Yamamoto et al., 2020, 2021). Moreover, by training with pre-trained CNN architectures, we hope to address the problem of small data size. We will opt to select the architectures that tallied the highest effect size on the AUC scores when the key clinical variables were integrated with the image-only models (Sukegawa et al. 2022; Yamamoto et al. 2020; Yamamoto et al. 2021) as detailed below:

##### ResNet50 architecture

ResNet50 has 50 layers that start with 7×7 convolutions followed by 3×3 convolutions in residual blocks. This architecture is known for its adaptability and robustness in medical imaging (Ho et al., 2021; Yamamoto et al., 2020; Yamamoto et al. 2021).

##### EfficientNetb3 architecture

This architecture was highlighted by its compound scaling method, which uniformly scales depth, width, and resolution, leading to improved accuracy and efficiency. With fewer parameters and accelerated processing capabilities, EfficientNetb3 is for medical imaging where dataset sizes can be limited (Sukegawa et al., 2022; Yamamoto et al., 2020).

To adapt these models to our task, we implemented transfer learning by initializing them with weights pre-trained on the ChestX-ray14 dataset, a large public dataset of chest radiographs (Cohen et al., 2021). We froze the early layers of each architecture responsible for capturing general image features such as edges and textures while unfreezing and fine-tuning the later layers to learn osteoporosis-specific patterns from our dataset. Additionally, the final fully connected classification layers were replaced with new layers compatible with our three-class output: normal, osteopenia, and osteoporosis. This approach allowed the models to retain foundational visual representations while adapting to the specific task of bone density classification.

#### Hyperparameter Experimentation

Hyperparameter experimentation was conducted to fine-tune the performance of both ResNet50 and EfficientNetB3 architectures. Hyperparameters are critical in guiding model behavior even before actual learning occurs, and the choice of settings affects convergence or cause overfitting. However, as there is no hyperparameter setting that applies for every algorithm, experimentation was necessary (Bergstra & Bengio 2012).

In this study, Optuna, a hyperparameter optimization framework, was used to automate the tuning process (*Optuna | Proceedings of the 25th ACM SIGKDD International Conference on Knowledge Discovery & Data Mining*, n.d.). Specifically, we used its Bayesian Optimization backend with a Median Pruner to efficiently terminate underperforming trials. Each trial aimed to maximize validation accuracy by sampling different combinations of three major hyperparameters: the number of epochs (10, 50, 100), learning rate (0.01, 0.05, 0.1), and batch size (4, 8, 16), as shown in Table 4.

**Table 4.**
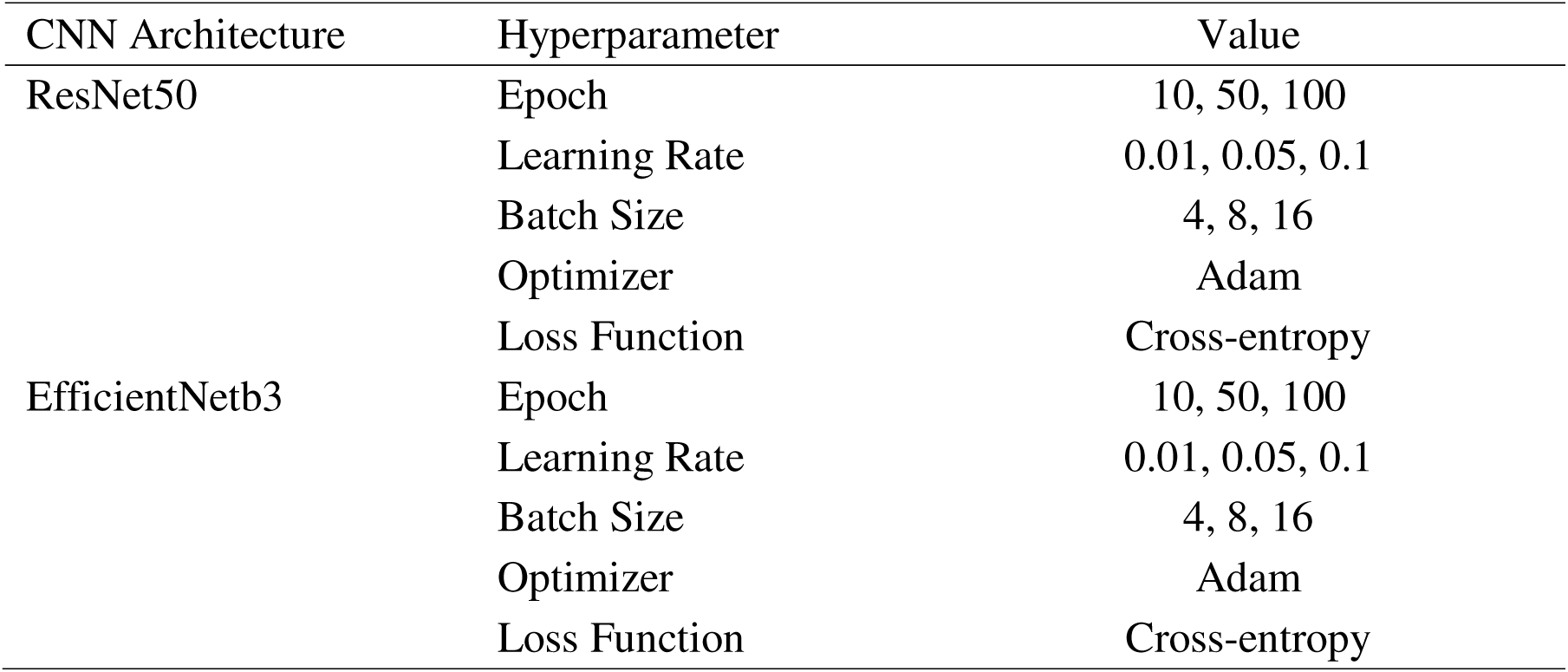
Hyperparameter Settings for each deep learning model.

Other training settings were kept constant across all experiments to ensure consistency and isolate the effects of hyperparameter tuning. We used the Adam optimizer for its adaptive learning rate and computational efficiency, along with a weight decay of 1×10^−4^to regularize the model and prevent overfitting.

The CrossEntropy loss function was selected as it is well-suited for multi-class classification problems. A StepLR scheduler was applied, reducing the learning rate by a factor of 0.5 every 5 epochs to facilitate better convergence as training progressed.

We employed Optuna, an automated framework that efficiently explores the hyperparameter search space. The objective function defined for Optuna dynamically instantiated the model and data loaders using each trial’s sampled parameters, such as batch size and learning rate, and trained the model on the training set. The function returned the validation loss as the metric to minimize, effectively guiding the optimization toward configurations that generalized well.

To avoid overfitting and unnecessary computational cost, we implemented early stopping with a patience of 20 epochs. That is, if the validation loss did not improve for 20 consecutive epochs, training was halted early. This strategy allowed us to terminate unpromising trials without compromising model performance.

A total of 10 Optuna trials were conducted per model architecture. The best-performing configuration from each run is reported in a summary table, streamlining the search for optimal training dynamics without extensive manual tuning.

### 3.7 Integrating Patient Variables to the Model

Two key fusion strategies (Pei et al. 2023) were deployed for this study, these are detailed below.

#### Early Fusion

The preprocessed X-ray images and the combined 1 × 3-dimensional vector was reshaped and embedded directly into the image tensor as additional pixel rows, slightly increasing the image height while preserving its single-channel format. This early fusion strategy allowed both ResNet50 and EfficientNetB3 to treat the clinical data as part of the visual input, enabling joint feature extraction from both modalities starting at the initial convolutional layers, as illustrated in (see Figure 5a). By incorporating clinical attributes directly into the image space, the models were able to learn spatially aware associations between patient characteristics and radiographic patterns. The resulting features were then passed through fully connected layers with ReLU activation to perform the final classification.

**Figure 5.**
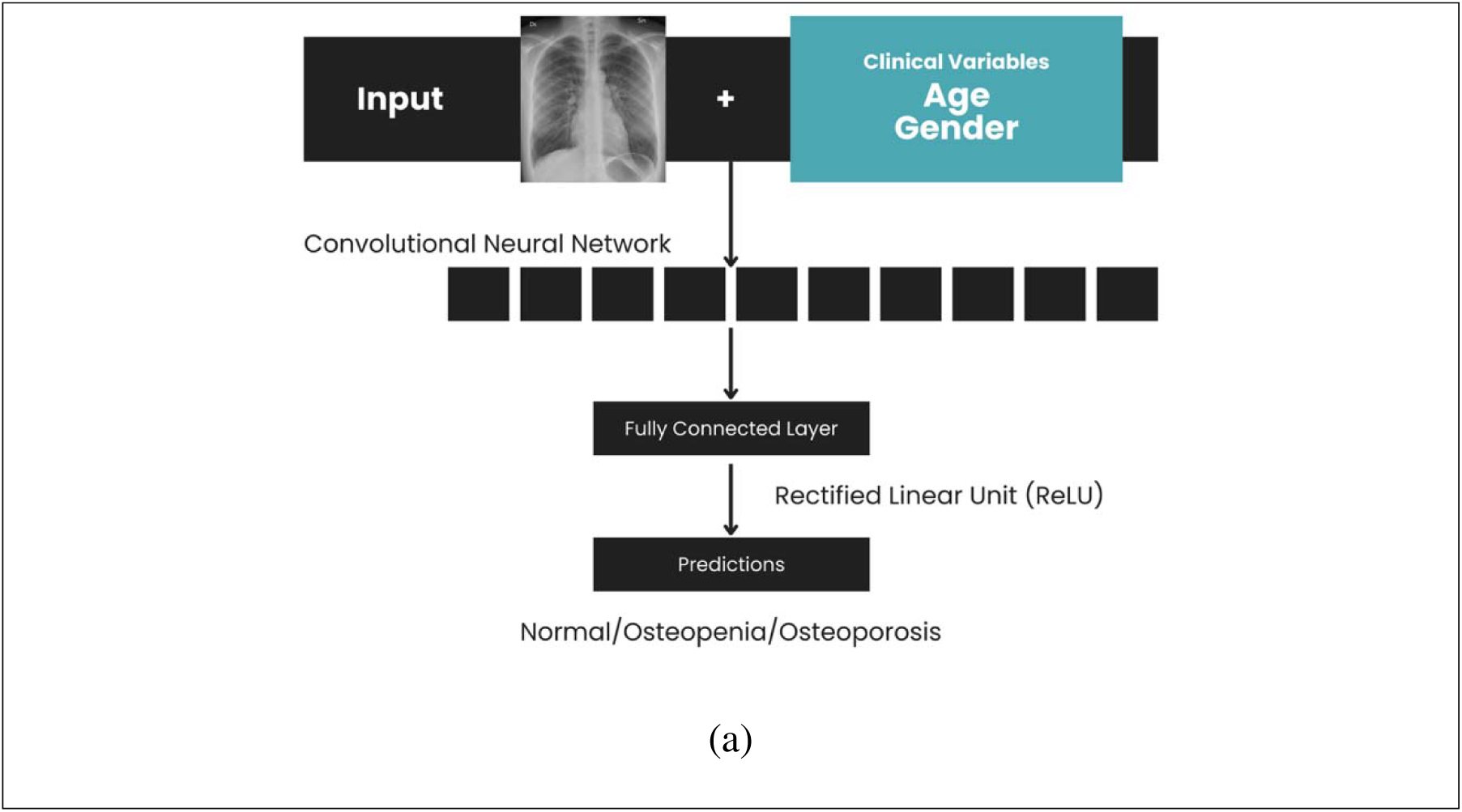
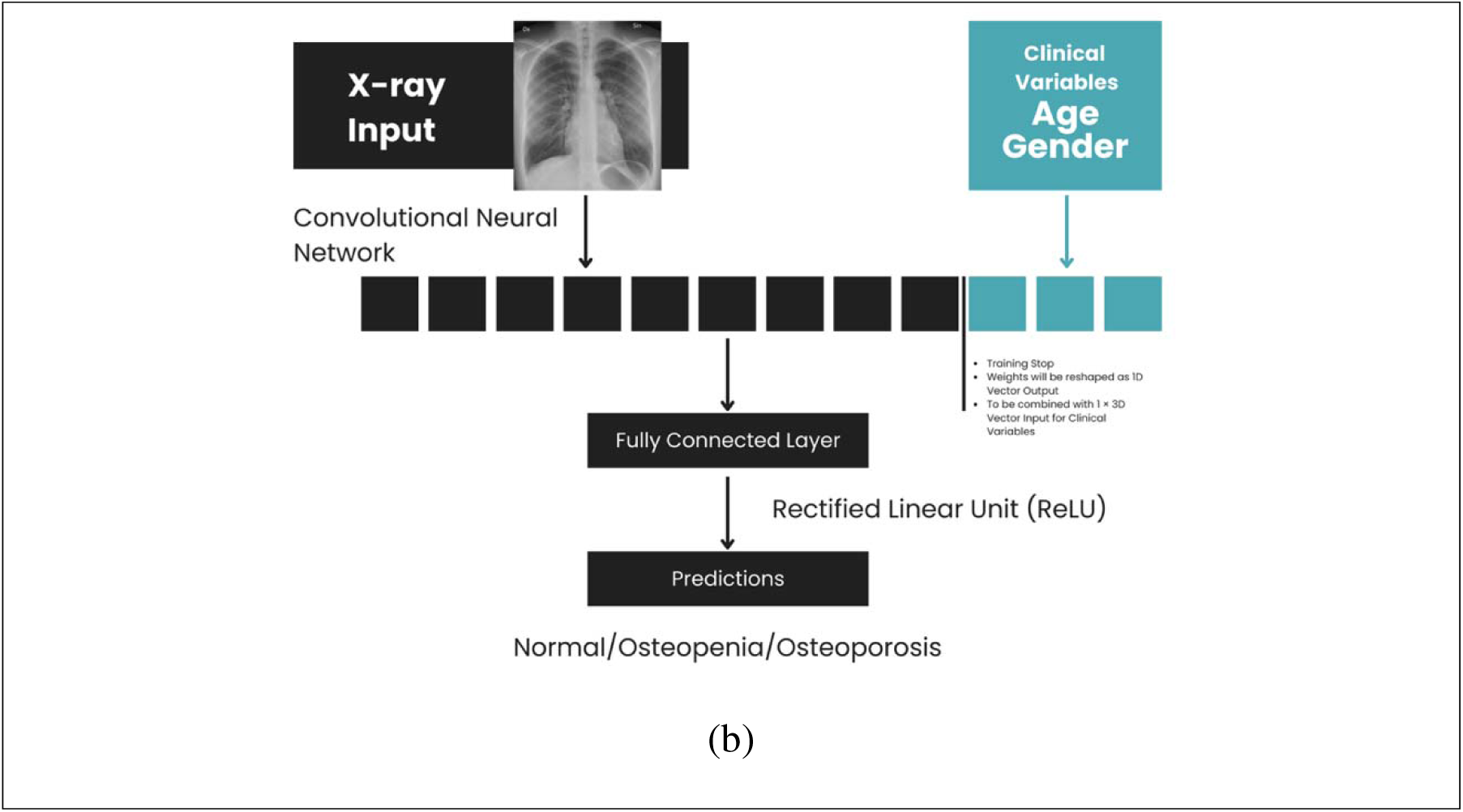
(a) Early Fusion Model for Osteoporosis Screening and (b) Late Fusion Model for Osteoporosis Screening

#### Late Fusion

The image data were first passed through the convolutional layers of the ResNet50 and EfficientNetB3 models to extract deep visual features, which were then reshaped into a one-dimensional vector. This feature vector (1536-dimensional) was concatenated with a 1 × 3-dimensional vector of preprocessed clinical variables, e.g., normalized age, BMI, and encoded gender, forming a unified multimodal representation (see Figure 5b). Instead of using the default classification head, we designed a custom Residual MLP Classifier, which took this concatenated vector as input. The MLP consisted of fully connected layers, including intermediate representations and residual connections, designed to enhance feature learning and classification. The final prediction was produced through this MLP, with the ReLU activation function applied within its hidden layers.

### 3.8 Model Comparison metrics

The models were assessed using several evaluation metrics derived from the confusion matrix: accuracy [6], precision [7], recall [8], specificity [9], sensitivity [10], F1 score [11], and the Area Under the Curve (AUC) score. These metrics provided a comprehensive overview of each model’s predictive performance.

Moreover, given that this is a three-class classification problem, the tabular depiction of actual vs predicted classification consists of the following,

True Positive (TP) : The number of correctly predicted positive cases for each class.

True Negative (TN) : The number of correctly predicted negative cases for each class.

False Positive (FP) : The number of negative cases incorrectly predicted as positive for each class.

False Negative (FN) : The number of positive cases incorrectly predicted as negative for each class.

We then computed the following metrics for each of the classes using the one-versus-all approach;

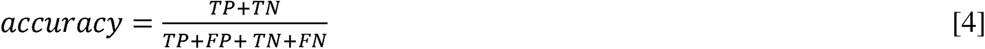

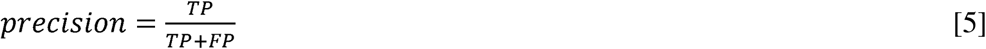

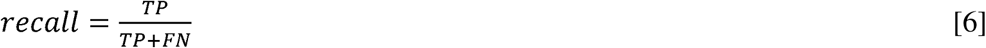

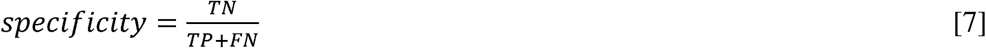

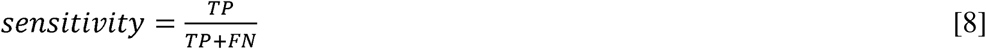

The macro-averaged F1-score was then computed by averaging the F1-scores of all three classes, as represented by equation [9]

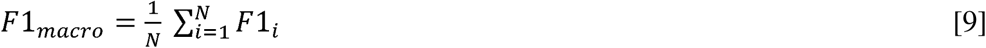

where,

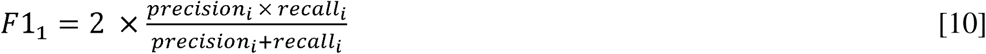

We then computed the following metrics for the overall performance of the model;

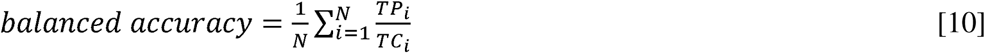

where,

*N* is the number of classes

*TP* is the number of correct predictions for class

*TC* is the number of actual instances of class

For computing the AUC score, we used the Weighted-average AUC approach where we computed a weighted average based on the number of true instances for each class. This method is appropriate given the imbalanced nature of our dataset. The Weighted AUC is given by [13].

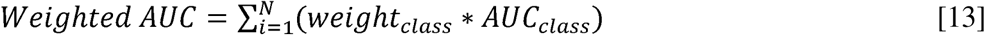

where,

*N* is the number of classes

*weigth_class_* is the proportion of the total sample belonging to class

*AUC_class_* is the computed AUC score for class

To ensure the reliability and validity of the model’s performance in real-world scenarios, we used the remaining 10% of the data that was allocated for testing. This portion of the dataset was not involved in the training or validation phases, thereby providing an unbiased assessment of the model.

### 3.9 Grad-CAM Visualization

We applied Grad-CAM to visualize the regions of the chest X-ray that significantly contributed to the predictions made by the image-only model in screening for osteoporosis. This provided clear visual representations through heat maps of the critical areas that influenced the model’s decision-making process.

These visualizations enhance model interpretability by highlighting image regions that the model found relevant for classification, allowing radiologists to validate whether these align with clinically meaningful anatomical structures. In this way, Grad-CAM bridges the gap between deep learning outputs and human interpretability, offering valuable insights for both diagnostic confidence and model trustworthiness.

### 3.10 Hardware and Software Specifications

All deep learning models were developed using Python and the PyTorch framework for model construction and optimization. Hyperparameter experimentation was conducted using Optuna, a robust tool that employs Bayesian optimization to efficiently explore the hyperparameter space and identify optimal configurations. The computational tasks were performed on an Acer Nitro 5 laptop equipped with an AMD Ryzen 5 5600H processor, NVIDIA GeForce RTX 3060 GPU with 6 GB of GDDR6 VRAM, 16 GB of DDR4 RAM, and a 512 GB SSD.

## 4 RESULTS AND DISCUSSION

This section presents the summary of the dataset and patient characteristics, followed by the evaluation of image-only and multimodal models across different Regions of Interest (ROIs). It also includes comparative benchmarking, model rankings, and Grad-CAM visualizations to support interpretability.

Patient demographics and clinical characteristics in the data are summarized in Table 5.

**Table 5.**
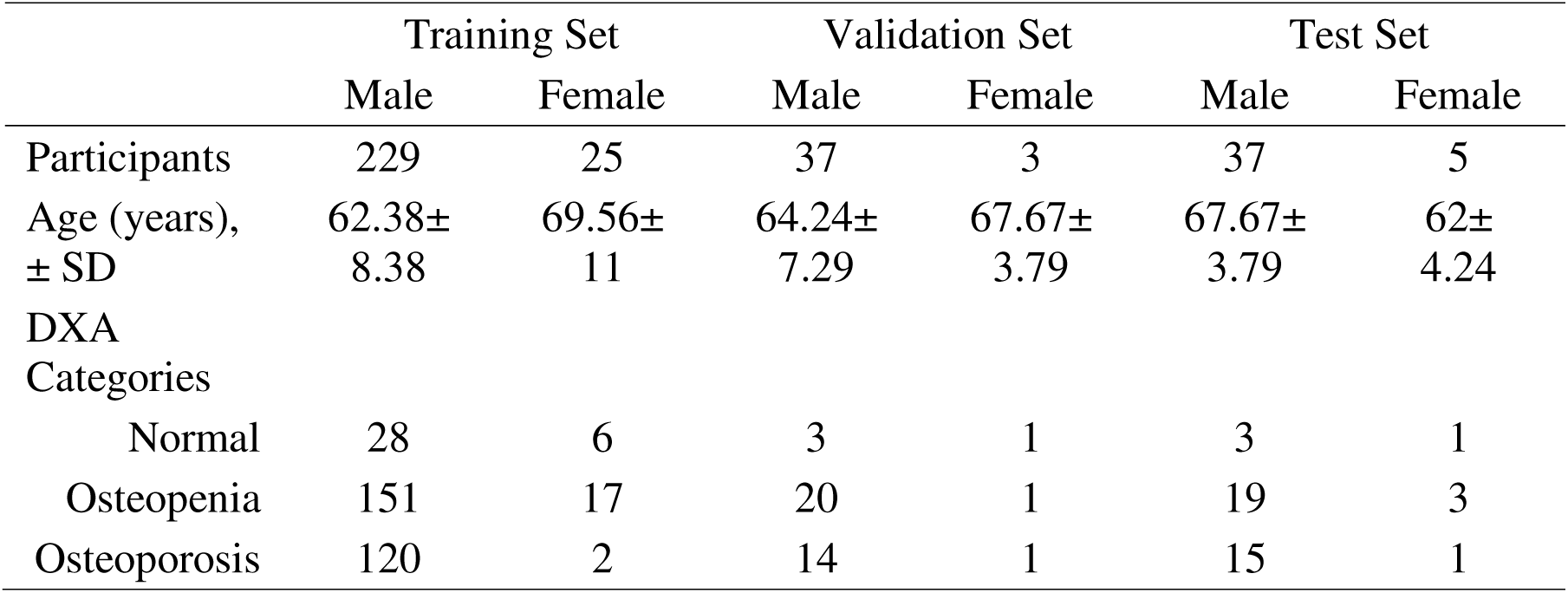
Summary of Participant Characteristics and DXA-Based Category Distribution Across the Training, Validation, and Test Sets.

|  | Training Set |  | Validation Set |  | Test Set |  |
| --- | --- | --- | --- | --- | --- | --- |
|  | Male | Female | Male | Female | Male | Female |
| Participants | 229 | 25 | 37 | 3 | 37 | 5 |
| Age (years),<br>± SD | 62.38±<br>8.38 | 69.56±<br>11 | 64.24±<br>7.29 | 67.67±<br>3.79 | 67.67±<br>3.79 | 62±<br>4.24 |
| DXA<br>Categories |  |  |  |  |  |  |
| Normal | 28 | 6 | 3 | 1 | 3 | 1 |
| Osteopenia | 151 | 17 | 20 | 1 | 19 | 3 |
| Osteoporosis | 120 | 2 | 14 | 1 | 15 | 1 |

**Table 6.**
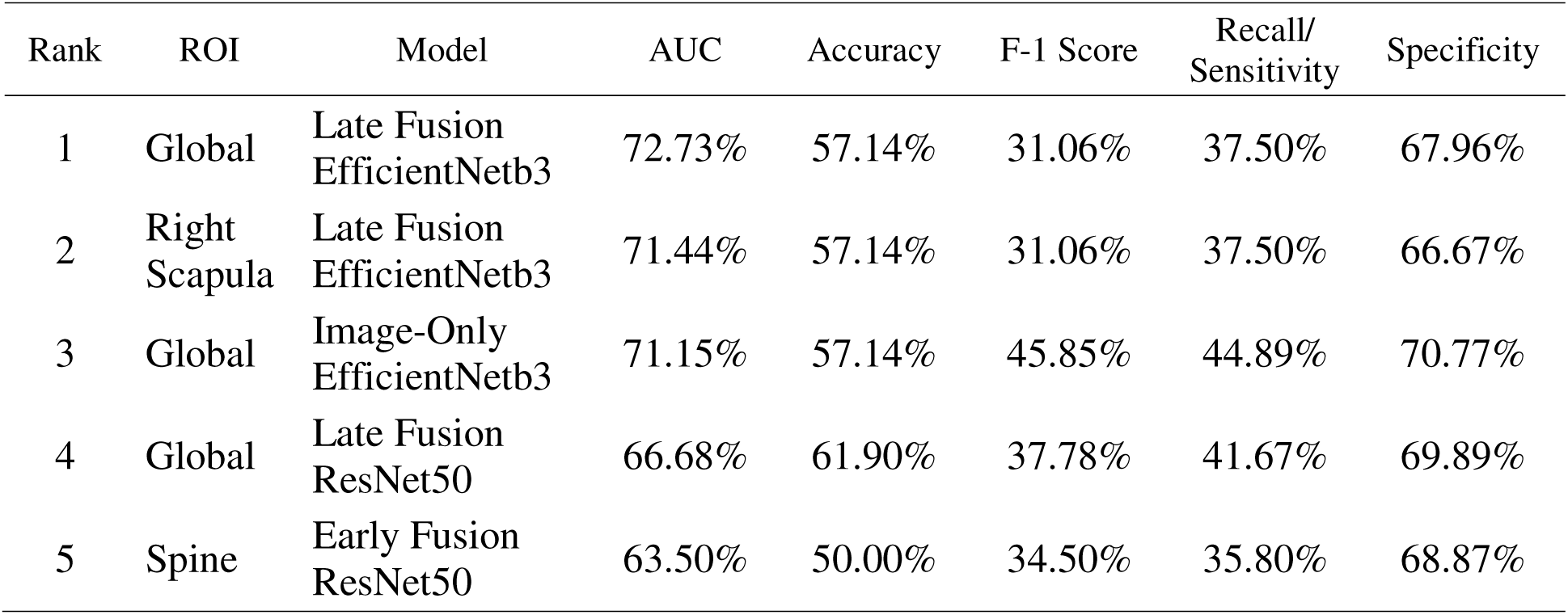
Ranked Summary of the Top Performing Models Across All ROIs and Modalities, Sorted by AUC.

The patient cohort predominantly comprised older adults, with mean ages ranging from 62 to 70 years across diagnostic categories. The dataset showed a notable class imbalance: osteopenia and osteoporosis cases accounted for 47.3% (n = 151) and 37.6% (n = 120) of the training set, respectively, while normal cases comprised only 8.8% (n = 28). The imbalance was reflected across all subsets and directly affected model calibration and performance, particularly precision and recall evaluation metrics.

While clinical literature identifies advancing age and female gender as key risk factors for osteoporosis, our dataset contained a relatively higher proportion of male patients across all classes. This deviation from expected clinical trends may reflect sampling characteristics of the archival dataset used, rather than actual population distributions. Statistical analysis further revealed significant differences in both age and gender across diagnostic groups (p < 0.001), reinforcing their potential discriminative power in model development. The DXA T-scores provided the reference standard for diagnosis, with the following thresholds applied: normal (T ≥ –1.0), osteopenia (–2.5 < T < –1.0), and osteoporosis (T ≤ –2.5).

### 4.1 CLAHE+Gamma Level Experimentation

To enhance the visibility of bone-related features crucial for osteoporosis detection, experiments were conducted to perform the most effective gamma correction level. Presented in Figure 6 were the images resulting from the CLAHE + Gamma Correction.

**Figure 6.**
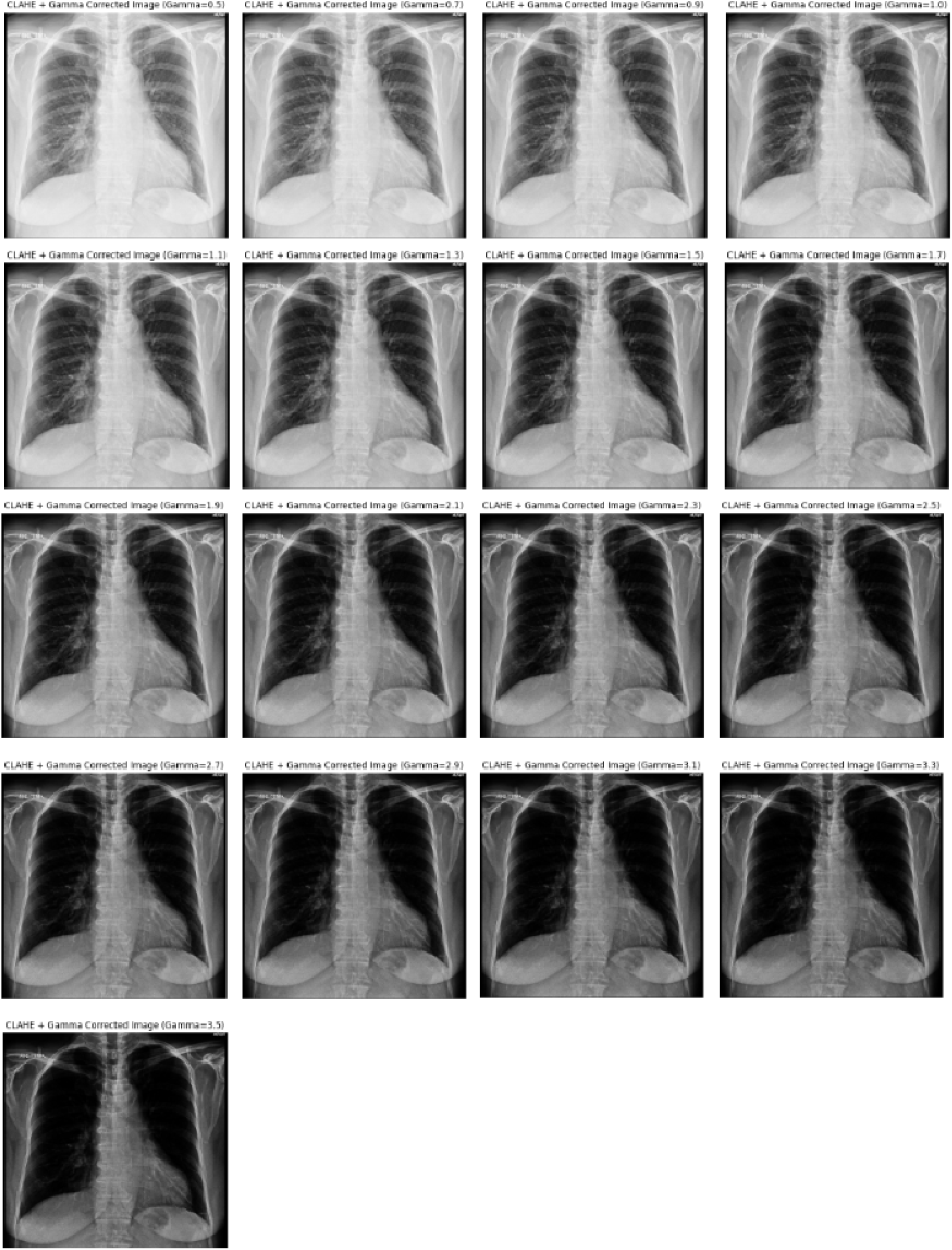
CLAHE + Gamma Correction Results.

For gamma values between 0.5 and 1.0, the images appeared significantly brighter, reducing contrast in high-intensity regions. This overexposure resulted in a loss of finer details in the cortical bone and trabecular structures of the vertebrae, which are crucial for osteoporosis assessment. Moreover, the increased brightness also diminished the visibility of rib contours and vertebral endplates, potentially affecting diagnostic accuracy.

Meanwhile, between gamma values of 1.1 and 1.9, the images exhibited a more balanced contrast enhancement, improving the distinction of mid-tone regions while preserving critical anatomical structures. The trabecular bone patterns within the vertebral bodies were more pronounced. Additionally, the costophrenic angles and rib structures were more clearly defined without significant noise amplification. The cortical bone margins of the vertebrae and clavicle

Lastly, for gamma values ranging from 2.1 to 3.5, the images appeared noticeably darker, which obscured low-intensity details but enhanced high-contrast structures. This adjustment improved the definition of cortical bone outlines, making vertebral fractures and variations in bone density more discernible. The posterior ribs and thoracic spine also exhibited higher contrast, which may assist in detecting subtle fractures.

The EfficientNetB3 (CLAHE + Gamma (γ = 1.7)) model demonstrated greater capability in class discrimination, particularly in identifying osteopenia and osteoporosis cases with fewer misclassifications compared to ResNet50.

Since all hyperparameters were held constant across both models, the observed performance differences can be attributed to the preprocessing technique. The CLAHE + Gamma configuration enhanced the radiographic features crucial for classification, enabling the model to better capture subtle visual cues. This led to improved balance in the prediction outcomes and a reduction in both false positives and false negatives.

#### Hyperparameter Experimentation, Model Training, Validation and Evaluation Results

To optimize model performance for each model, including ResNet50 and EfficientNetB3 under Image-Only, Early Fusion and Late Fusion approaches, the hyperparameter experimentation was performed separately for each model. While the discussed set-up was consistent, the optimal hyperparameters may vary depending on the architecture and fusion strategy as each model has different convergence behavior and sensitivity to hyperparameter adjustments.

#### Global Region of Interest (ROI)

Each model underwent a dedicated Optuna-guided hyperparameter search, tuning key parameters such as batch size, learning rate, and number of training epochs.

Among all models evaluated, the Late Fusion ResNet50 yielded the highest validation accuracy at 72.50%, followed by Early Fusion ResNet50 at 70.00%. Notably, configurations with lower learning rates (0.01) paired with moderate to large batch sizes (≥ 8) frequently led to more stable convergence, particularly for EfficientNetB3. In contrast, ResNet50 generally benefitted from slightly higher learning rates (0.05) and smaller batch sizes, especially under early fusion. These results underscore the importance of model-specific hyperparameter tuning, particularly in multimodal settings where data integration strategy significantly affects learning dynamics and generalization.

Among all configurations, Late Fusion ResNet50 yielded the highest accuracy at 61.90%, followed by Late Fusion EfficientNetB3 and Image-Only EfficientNetB3, both at 57.14%. The consistent top-tier performance of the late fusion models highlights the benefit of integrating clinical covariates at the feature level, where structured variables are combined with learned image representations after convolution.

In terms of AUC, which reflects a model’s ability to distinguish between classes, Late Fusion EfficientNetB3 outperformed all others with a score of 72.73%. The Image-Only EfficientNetB3 model closely followed with an AUC of 71.15%, indicating that this architecture remains highly effective even without multimodal integration. By contrast, the ResNet50 counterparts, particularly the Image-Only model, lagged behind with substantially lower AUC (52.89%) and accuracy (40.48%), reinforcing the superior representational power of EfficientNetB3 in radiographic tasks.

Precision and F1 Score were highest in Image-Only EfficientNetB3 (71.85% and 45.85%, respectively), though its recall/sensitivity was relatively low at 44.89%, suggesting a tendency to favor confident positive predictions while missing some true positives. In comparison, Late Fusion EfficientNetB3 delivered a more balanced performance with strong scores across all key metrics, supported by a competitive precision of 51.67%, recall/sensitivity of 37.50%, and an F1 Score of 31.06%.

Performance in terms of recall/sensitivity, which is the model’s capacity to correctly identify true positive cases was highest for Late Fusion ResNet50 (41.67%), followed by Early Fusion EfficientNetB3 (39.58%). This suggests that models leveraging late and early fusion were more responsive to osteoporotic indicators embedded in both image and clinical data. On the other hand, specificity, or the ability to correctly identify non-osteoporotic cases, peaked at 72.54% in Image-Only ResNet50 and remained high in Image-Only EfficientNetB3 (70.77%) and Late Fusion ResNet50 (69.89%), reflecting strong negative class discrimination.

Early Fusion approaches were consistently outperformed by late fusion and image-only baselines. Both ResNet50 and EfficientNetB3 exhibited reduced classification performance when clinical covariates were concatenated at the pixel level, potentially due to the distortion of spatial patterns within the input tensors. This was particularly evident in Early Fusion ResNet50, which yielded the lowest F1 Score (29.79%) and precision (28.91%).

These findings affirm that Late Fusion, particularly when implemented with EfficientNetB3, provides the most robust and diagnostically balanced architecture in this study. The approach not only outperformed early fusion strategies but also delivered meaningful improvements over image-only baselines, highlighting the advantage of feature-level integration in multimodal osteoporosis screening.

However, it is important to contextualize these results within the inherent class imbalance of the dataset, where Osteopenia constituted the majority class while Normal and Osteoporosis cases were relatively underrepresented. This imbalance likely contributed to the skewed recall and precision scores observed across several models, especially those that consistently misclassified Normal cases or overpredicted Osteopenia.

The confusion matrices provide insight into how each model distributes its predictions across the three classes and reveal patterns that complement the metric-based evaluation. The ResNet50 Image-Only model exhibited substantial misclassifications, with Osteoporosis and Normal cases frequently predicted as Osteopenia. Its limited ability to detect true positive cases, particularly in the Osteoporosis class, is reflected in its low sensitivity and F1 Score.

The ResNet50 Early Fusion model showed slight improvement in Osteopenia identification but continued to misclassify a majority of Osteoporosis cases. The model predicted no Normal instances correctly, suggesting limited generalization at the pixel-level fusion stage. This performance pattern underscores the disruption in spatial coherence that may arise from early fusion using clinical covariates.

In contrast, the ResNet50 Late Fusion model demonstrated a more focused prediction profile, correctly identifying 22 Osteopenia and 4 Osteoporosis cases. Although it still failed to predict any Normal cases, its overall structure of predictions suggests that feature-level fusion allowed for more nuanced learning, especially in distinguishing Osteopenia from Osteoporosis.

Turning to EfficientNetB3, the Image-Only model exhibited stronger class-wise prediction accuracy, correctly identifying 20 Osteopenia and 3 Osteoporosis cases. The precision and recall advantages previously noted are clearly reflected in its relatively well-distributed predictions, even in the absence of clinical data.

The EfficientNetB3 Early Fusion model achieved more balanced classification, correctly predicting 11 cases each for Osteopenia and Osteoporosis. However, consistent with other early fusion approaches, it struggled with the Normal class. This further suggests that while EfficientNetB3’s architecture is more resilient, pixel-level fusion still introduces noise that affects early convolutional representations.

Finally, the EfficientNetB3 Late Fusion model produced the most structured and clinically reliable confusion matrix. It achieved 22 correct predictions for Osteopenia and 14 for Osteoporosis, reflecting strong performance on the two clinically significant classes. Although Normal cases remained underclassified, this can be partially attributed to both the data imbalance and the model’s optimization towards the majority and more visually complex classes. The matrix supports the model’s high AUC and balanced sensitivity-specificity tradeoff, making it the most reliable architecture in this study.

#### Right Clavicle ROI

Among all configurations, both Late Fusion ResNet50 and Late Fusion EfficientNetB3 achieved the highest validation accuracy at 62.50%, followed closely by Early Fusion EfficientNetB3 at 60.01%. A closer look at their hyperparameter settings reveals key trends. For instance, the Late Fusion models performed best when trained with a batch size of 16, suggesting that larger batch updates may be more effective for multimodal data integration in localized ROIs. Additionally, the learning rate of 0.01 for ResNet50 and 0.05 for EfficientNetB3 points to architecture-specific sensitivity, where more aggressive learning was better tolerated by EfficientNetB3, possibly due to its compound scaling design.

Both early fusion models converged under higher learning rates (0.1) and smaller batch sizes (4), a combination that typically encourages rapid learning but can risk instability. That they still achieved strong validation performance suggests that early fusion benefitted from more fine-grained updates, especially when fusing low-level features. Meanwhile, the image-only models, despite using moderate learning rates (0.05 and 0.01), lagged behind in accuracy. This performance gap reinforces the importance of clinical covariates in enhancing discriminative capacity for this ROI, especially under imbalanced class scenarios and subtle radiographic differences.

Late Fusion ResNet50 achieved the highest accuracy (52.38%), followed closely by Late Fusion EfficientNetB3 and Image-Only EfficientNetB3 (both at 50.00%). These results suggest that integrating clinical covariates at the feature level leads to modest improvements over image-only configurations for this region.

In terms of AUC, Late Fusion EfficientNetB3 recorded the highest score (62.08%), followed by Early Fusion EfficientNetB3 (60.58%), reflecting the consistent advantage of EfficientNetB3 when clinical data is introduced. Notably, Early Fusion EfficientNetB3 also obtained the highest recall/sensitivity (49.62%) and F1 score (39.23%), indicating its strength in detecting positive cases. This is particularly relevant in clinical settings where minimizing false negatives is crucial.

Image-Only ResNet50 performed the weakest across all metrics, reaffirming its limitations in both unimodal and multimodal settings. In contrast, Image-Only EfficientNetB3 remained competitive, especially in terms of precision (40.50%) and AUC (56.07%), reinforcing its strength as a standalone image-based classifier.

The ResNet50 Image-Only model frequently misclassified Normal and Osteoporosis cases as Osteopenia, with no correct predictions for Osteoporosis. Similarly, ResNet50 Early Fusion improved slightly in classifying Osteopenia but continued to misclassify the other classes. Late Fusion ResNet50 showed clearer improvements, correctly identifying 19 Osteopenia and 3 Osteoporosis cases. However, like the previous configurations, it failed to predict any Normal cases correctly.

EfficientNetB3-based models demonstrated more balanced performance. Image-Only EfficientNetB3 effectively identified Osteopenia and Osteoporosis cases but misclassified all Normal cases. Early Fusion EfficientNetB3, however, achieved correct predictions across all three classes, with 3 Normal, 8 Osteopenia, and 6 Osteoporosis cases correctly identified. This reflects its high recall and F1 score in the earlier metrics. Late Fusion EfficientNetB3 also performed well, correctly identifying 17 Osteopenia and 4 Osteoporosis cases, although Normal cases were again missed.

EfficientNetB3 consistently outperformed ResNet50 across all fusion strategies. While Late Fusion models achieved higher accuracy, Early Fusion with EfficientNetB3 stood out for its recall and class-wise balance, showing potential in sensitivity-driven applications. The persistent underperformance in recognizing Normal cases across all models likely reflects the influence of class imbalance within the dataset.

#### Right Scapula ROI

As part of the model optimization process for the Right Scapula Region of Interest (ROI), each model architecture underwent independent hyperparameter tuning using Optuna. This process involved varying combinations of batch size, learning rate, and epoch count to maximize validation accuracy specific to each model setup, whether image-only, early fusion, or late fusion.

Both Early Fusion ResNet50 and Early Fusion EfficientNetB3 achieved the top performance, each recording a validation accuracy of 70.01%, suggesting that early integration of clinical covariates is particularly effective in extracting meaningful features from scapular images. In comparison, Late Fusion ResNet50 followed closely with 65.00%, while the Image-Only variants lagged behind, with validation scores below 50%.

A key trend observed in the best-performing configurations was the use of larger batch sizes (16) in early fusion models, indicating that processing more data per update stabilized training during multimodal learning. In contrast, late fusion models favored smaller batch sizes (4), likely due to the localized impact of appended clinical covariate pixels, which required finer gradient updates for convergence. Across all models, a learning rate of 0.01 was consistently selected, reinforcing its reliability in achieving smooth optimization within this ROI.

Image-Only EfficientNetB3 achieved the highest accuracy at 59.52%, followed by Late Fusion EfficientNetB3 (57.14%) and Early Fusion EfficientNetB3 (54.76%). These results suggest that EfficientNetB3 continues to deliver stable performance on this local ROI regardless of fusion strategy, with image-only and fusion-based models both achieving competitive results.

When considering AUC as a measure of discriminatory ability, Late Fusion EfficientNetB3 stood out with the highest score (71.44%), further reinforcing its ability to distinguish between classes effectively. This was followed by Image-Only EfficientNetB3 (61.88%) and Early Fusion EfficientNetB3 (60.51%), again showing consistent superiority over ResNet50-based models.

Early Fusion EfficientNetB3 achieved the highest F1 Score (49.38%) and the highest recall/sensitivity (51.33%), suggesting that pixel-level integration with clinical variables, when combined with a strong backbone, may improve the model’s ability to capture positive cases without significantly compromising precision. While Late Fusion EfficientNetB3 offered the best AUC and strong specificity (66.67%), Early Fusion appeared to favor recall, reflecting a similar trade-off observed in the Right Clavicle ROI. Meanwhile, ResNet50 models, particularly those using fusion strategies, underperformed. Late Fusion ResNet50 recorded the lowest accuracy (45.24%) and AUC (44.20%), with limited gains in recall or precision. Image-Only ResNet50 and Early Fusion ResNet50 showed similar patterns, further affirming the limitations of this backbone for the scapula region.

In contrast, EfficientNetB3 models demonstrated better class differentiation. Image-Only EfficientNetB3 correctly predicted 22 Osteopenia and 3 Osteoporosis cases, and Early Fusion EfficientNetB3 further improved this balance with correct predictions across all three classes: 2

Normal, 16 Osteopenia, and 5 Osteoporosis. Late Fusion EfficientNetB3 maintained high Osteopenia classification (22 correct) and correctly predicted 2 Osteoporosis cases, although Normal predictions remained absent.

These results reinforce EfficientNetB3’s consistent advantage across local ROIs, with Early Fusion excelling in sensitivity-related metrics and Late Fusion showing strength in AUC and specificity. The confusion matrices further highlight the effect of class imbalance, as Normal cases remained the least predicted across all configurations. Nonetheless, among all model types, EfficientNetB3 Early Fusion demonstrated the best overall class-wise balance for the Right Scapula, making it a strong candidate for sensitivity-oriented clinical screening.

#### Spine ROI

Each model trained under the Spine Region of Interest (ROI) underwent dedicated hyperparameter tuning using Optuna, optimizing combinations of batch size, learning rate, and training epochs specific to each fusion strategy—image-only, early fusion, or late fusion.

Among all models, Early Fusion ResNet50 achieved the highest validation accuracy at 72.50%, closely followed by Early Fusion EfficientNetB3 at 70.00%. These results reinforce the strength of early-stage clinical covariate integration in modeling spine-specific structural patterns associated with osteoporosis.

In contrast, Late Fusion ResNet50 recorded a solid 62.50% despite training for only 10 epochs, suggesting rapid convergence, while image-only models posted comparatively lower scores (56.53% and 57.87%), emphasizing the diagnostic limitations of relying on visual data alone.

A key trend across the best-performing setups was the use of batch sizes of 8 for early fusion models, which may have facilitated stable learning in the multimodal setting. Meanwhile, smaller batch sizes (4) were optimal for late fusion, where fine-tuning weight updates to accommodate pixel-level covariates was likely beneficial. Additionally, a learning rate of 0.01 was favored in most configurations, suggesting it provided reliable gradient descent behavior for this ROI.

Late Fusion ResNet50 achieved the highest accuracy (54.76%) among all configurations, followed by Late Fusion EfficientNetB3 (52.38%) and Image-Only EfficientNetB3 and Early Fusion ResNet50 (both at 50.00%). These results show a modest but notable performance advantage for fusion-based models, particularly when using the ResNet50 backbone, in contrast to trends observed in other ROIs.

In terms of AUC, Early Fusion ResNet50 surprisingly outperformed all other models with a score of 63.50%, indicating its relatively strong class-separation ability despite moderate accuracy. Early Fusion EfficientNetB3 followed with 59.81%, while Late Fusion ResNet50 recorded a lower AUC at 57.50% but still outperformed other ResNet50 variants.

In terms of recall/sensitivity and F1 Score, Late Fusion ResNet50 also led with values of 48.48% and 35.71%, respectively. These scores indicate its stronger capability to detect positive cases compared to other models, though it traded off some precision (28.62%). Meanwhile, Image-Only EfficientNetB3 recorded the highest precision (40.51%) but showed a reduced F1 score (34.50%) due to lower recall.

Image-Only ResNet50 demonstrated the weakest performance overall, with an extremely low accuracy (9.52%) and F1 Score (5.80%). The model struggled with both recall and precision, as also observed in the confusion matrix, where the majority of predictions were incorrectly concentrated in the Normal class regardless of the actual label.

The Image-Only ResNet50 model exhibited near-complete misclassification, assigning nearly all inputs to the Normal class. In contrast, Late Fusion ResNet50 showed more refined predictions, with better separation between Osteopenia and Osteoporosis cases, contributing to its higher recall.

EfficientNetB3 models, particularly the early and late fusion variants, showed class-wise improvements in recall over their image-only counterpart. Early Fusion EfficientNetB3 correctly predicted a mix of Osteopenia and Osteoporosis cases, showing a more even distribution. Late Fusion EfficientNetB3 had a more focused prediction on the Osteopenia class, which aligns with its better performance in specificity and AUC.

These findings suggest that while EfficientNetB3 remains a reliable architecture, the Spine ROI presents a more complex challenge where ResNet50—with late fusion—performed competitively. Compared to other local ROIs, the spine showed greater variability in performance across fusion strategies, emphasizing the importance of ROI-specific tuning in multimodal osteoporosis screening.

Full optimization and classification results, including detailed trial logs, validation performance per configuration, and confusion matrices are provided as supplementary materials. These supplementary outputs highlight the variability across trials and reinforce the final configurations summarized.

#### Top Performing Models and Comparative Benchmarking with Existing Studies

Table 5 ranks the top-performing models across all ROIs and fusion strategies based on the AUC score, which serves as the primary metric for evaluation the models’ overall discriminatory power.

One observation stood out, that is the late fusion approaches consistently outperform their early fusion and image-only counterparts, with EfficientNetb3 leading across multiple configurations. This reinforces the strength of combining structured clinical data at the feature-level, particularly when part the said CNN architecture.

As a matter of fact, the top three of the five models in Table 40 are Late Fusion EfficientNetb3 configurations–– two of which are Global and Right Scapula ROIs, ranked first and second, respectively. Their consistent AUC values above 71% in both global and local contexts highlight EfficientNetb3’s ability to generalize well and extract complementary information from multimodal sources.

Moreover, even the Image-Only EfficienNetb3 ranked third, surpassing all ResNet50-based approaches. The rankings suggest that EfficientNetb3’s feature extraction capabilities are robust enough to perform competitively even without the additional clinical variables. Its top-ranking F1 and sensitivity values also emphasize its strength in correctly identifying positive cases.

On the other hand, ResNet50-based models, while occasionally competitive in specific ROIs like the Spine, lagged in general performance. Their lower AUC and sensitivity values suggest limitations in capturing subtle clinical patterns, especially when early fusion is used. Although Early Fusion ResNet50 for the Spine ROI managed to break into the top five, its position underscores that such outcomes may be ROI-specific rather than consistent across the dataset.

The results underscore two core findings: first, that EfficientNetB3 is the most adaptable and high-performing backbone in this multimodal screening framework; and second, that late fusion offers a more effective integration strategy than early fusion, striking a better balance between image representation and clinical context. To assess how these results align with or diverge from prior work, we benchmarked the top models in this study against published approaches using other X-ray modalities.

As shown in Table 7, existing studies by Yamamoto et al. (2020, 2021) and Sukegawa et al. (2022) demonstrated high AUC scores ranging from 88.5% to 93.7% using hip and panoramic X-rays with ResNet50 and EfficientNetB3. These imaging modalities directly capture trabecular bone structures and offer rich visual cues, contributing to their superior model performance in osteoporosis detection. In contrast, this study employed chest X-rays, an unconventional and more complex imaging source for bone health assessment, yet still achieved an AUC of 72.73% using Late Fusion EfficientNetB3. This outcome highlights the feasibility of using chest radiographs for opportunistic osteoporosis screening, especially when traditional sites like the hip or spine are unavailable.

**Table 7.** Comparative Summary of AUC Scores from Prior Studies and This Study’s Multimodal Models.

| CNN Architecture | Author | Medical Images | Total Sample Size | Class Balance | AUC Score |  | Effect Size |
| --- | --- | --- | --- | --- | --- | --- | --- |
|  |  |  |  |  | Image-only | Ensemble |  |
| ResNet50 | Yamamoto et al. (2021) | Hip X-ray | 1,699 | Balanced | 88.5% | 90.6% | 2.1% |
|  | ***This study*** | ***Chest X-ray*** | ***408*** | ***Imbalanced*** | ***52.89%*** | ***66.68%*** | ***13.79%*** |
| EfficientNet b3 | Yamamoto et al. (2020) | Hip X-ray | 1,133 | Balanced | 90.9% | 93.7% | 2.8% |
|  | Sukegawa et al., (2022) | Panoramic X-ray | 778 | Imbalanced | 86.7% | 89.9% | 3.2% |
|  | ***This study*** | ***Chest X-ray*** | ***408*** | ***Imbalanced*** | ***71.15%*** | ***72.73%*** | ***1.58%*** |

Notably, the effect size improvements between image-only and multimodal models were especially significant for ResNet50, which rose by 13.79%, and modestly for EfficientNetB3 at 1.58%. This supports the notion that integrating clinical covariates is most beneficial in cases where the image modality provides less direct information, as is true for chest X-rays. The multimodal approach serves as a compensatory mechanism, enhancing prediction even when the imaging source is suboptimal.

Beyond imaging modality, class distribution may have played a crucial role in evaluation outcomes. Prior studies generally operated on balanced datasets (e.g., Yamamoto et al. with 598 vs. 535 and 909 vs. 790 class counts), which promote fair learning and reduce bias during training. In contrast, this study and Sukegawa et al. worked with imbalanced datasets, where the dominance of one class can lead to skewed predictions and underperformance in minority class recognition. This imbalance likely contributed to the lower absolute AUCs observed in this study, particularly in image-only configurations. However, the substantial effect size improvements seen, especially in the ResNet50 models, demonstrate how fusion with clinical variables can help mitigate the performance penalty associated with class imbalance.

Altogether, these findings underscore two key insights: first, chest X-rays, though anatomically less direct, can serve as a viable modality for osteoporosis screening when paired with clinical data; and second, multimodal approaches offer greater performance gains in settings with either low-information input (e.g., chest X-rays) or imbalanced datasets, reinforcing their value in real-world clinical applications.

#### GRAD-CAM Results

The Grad-CAM overlays in Table 8 visualize the regions attended to by the ResNet50 model during both correct and incorrect classifications of Normal and Osteopenia cases. These attention maps provide interpretability into how the model makes decisions and whether its focus aligns with anatomically relevant structures.

**Table 8.**
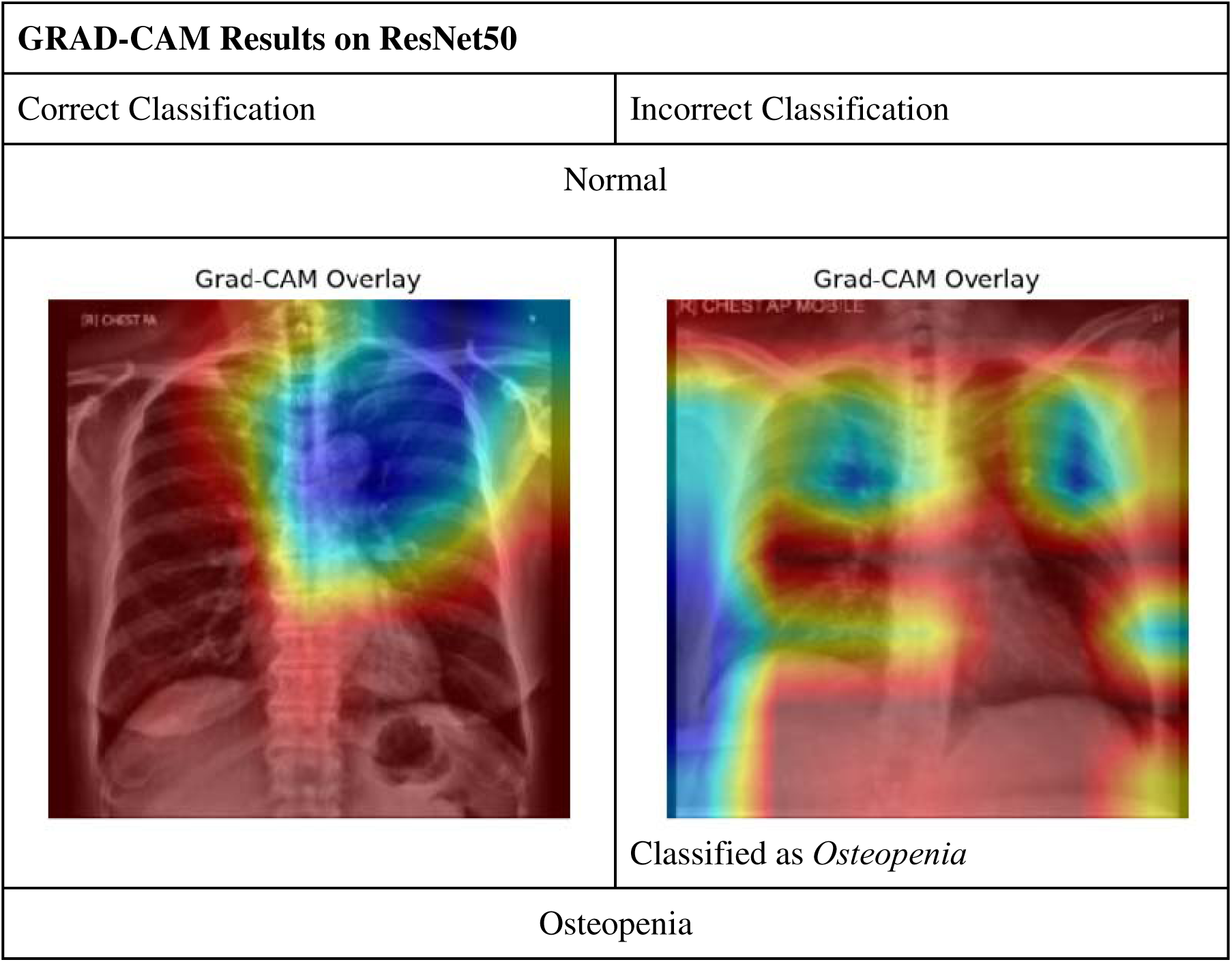
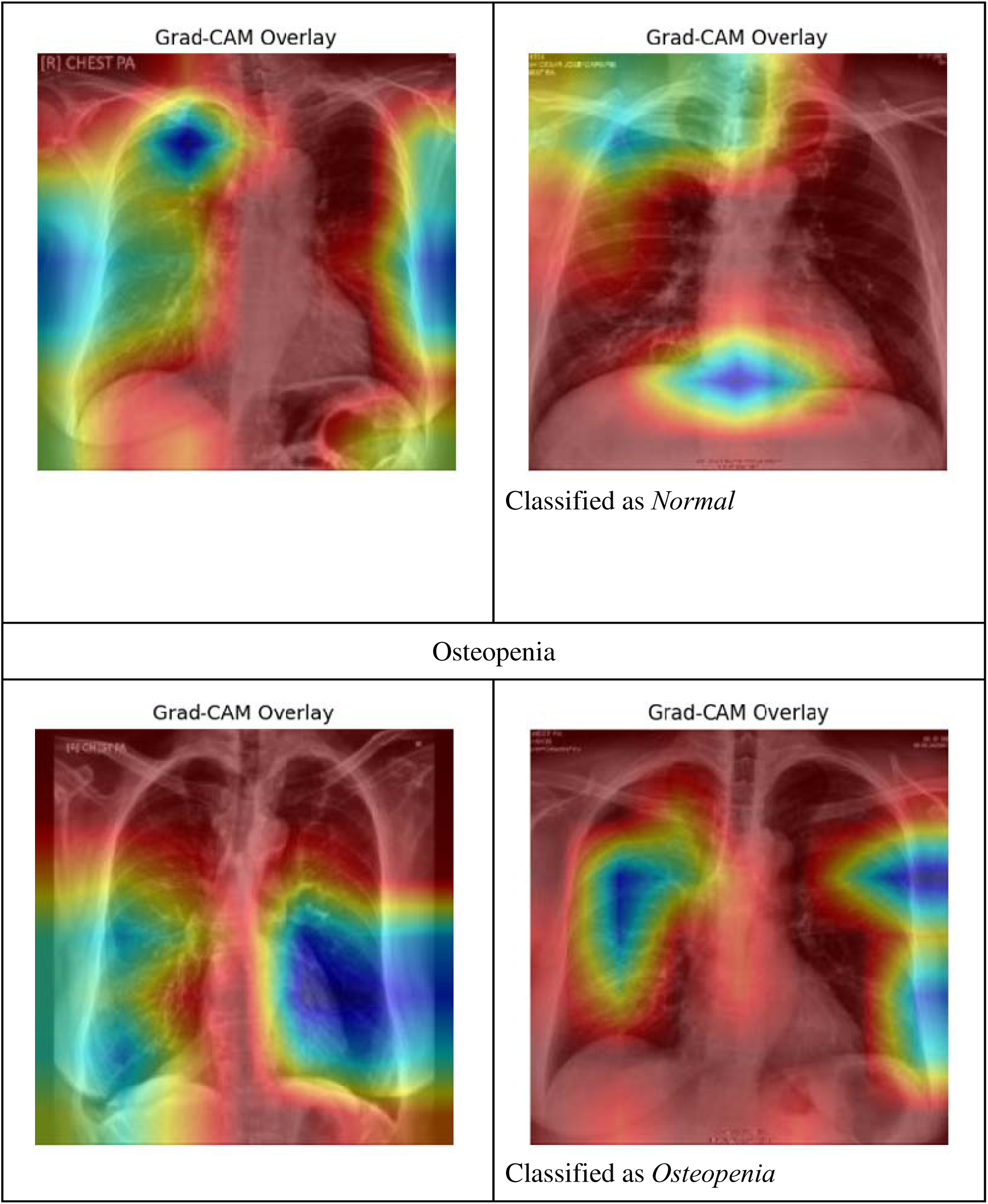
Comparison of Grad-CAM Results on the ResNet50 Model Across Correct and Incorrect Classifications.

| GRAD-CAM Results on ResNet50 |  |
| --- | --- |
| Correct Classification | Incorrect Classification |
| Normal |  |
| Grad-CAM Overlay | Grad-CAM Overlay<br>Classified as *Osteopenia* |
| Osteopenia |  |
| Grad-CAM Overlay | Grad-CAM Overlay<br>Classified as *Normal* |
| Osteopenia |  |
| Grad-CAM Overlay | Grad-CAM Overlay<br>Classified as *Osteopenia* |

In correctly classified samples, the model demonstrates focused attention on clinically significant regions. For Normal predictions, the model predominantly attends to the upper thoracic cavity, particularly around the clavicle and lung apex—regions commonly examined in osteoporosis screening. Similarly, in correctly classified Osteopenia cases, attention is concentrated on the scapular region and lower thoracic spine, suggesting that the model successfully captures relevant skeletal features when confident in its prediction.

Conversely, in misclassified instances, the attention becomes more diffused and erratic. Incorrect classifications of Normal cases reveal scattered attention across bilateral lung fields and soft tissue areas, indicating that the model may be relying on less informative or noisy features. Misclassified Osteopenia examples further illustrate this inconsistency, as the Grad-CAM highlights broad and non-discriminative areas, including central thoracic and mediastinal regions—areas less indicative of bone density variation.

Table 9 visualizes the Grad-CAM overlays for EfficientNetB3, showing both correct and incorrect predictions for Normal and Osteopenia cases. Compared to ResNet50, the attention maps generated by EfficientNetB3 appear more centralized, with greater focus on the vertebral column and lower thoracic region—key anatomical sites for osteoporosis-related changes.

**Table 9.**
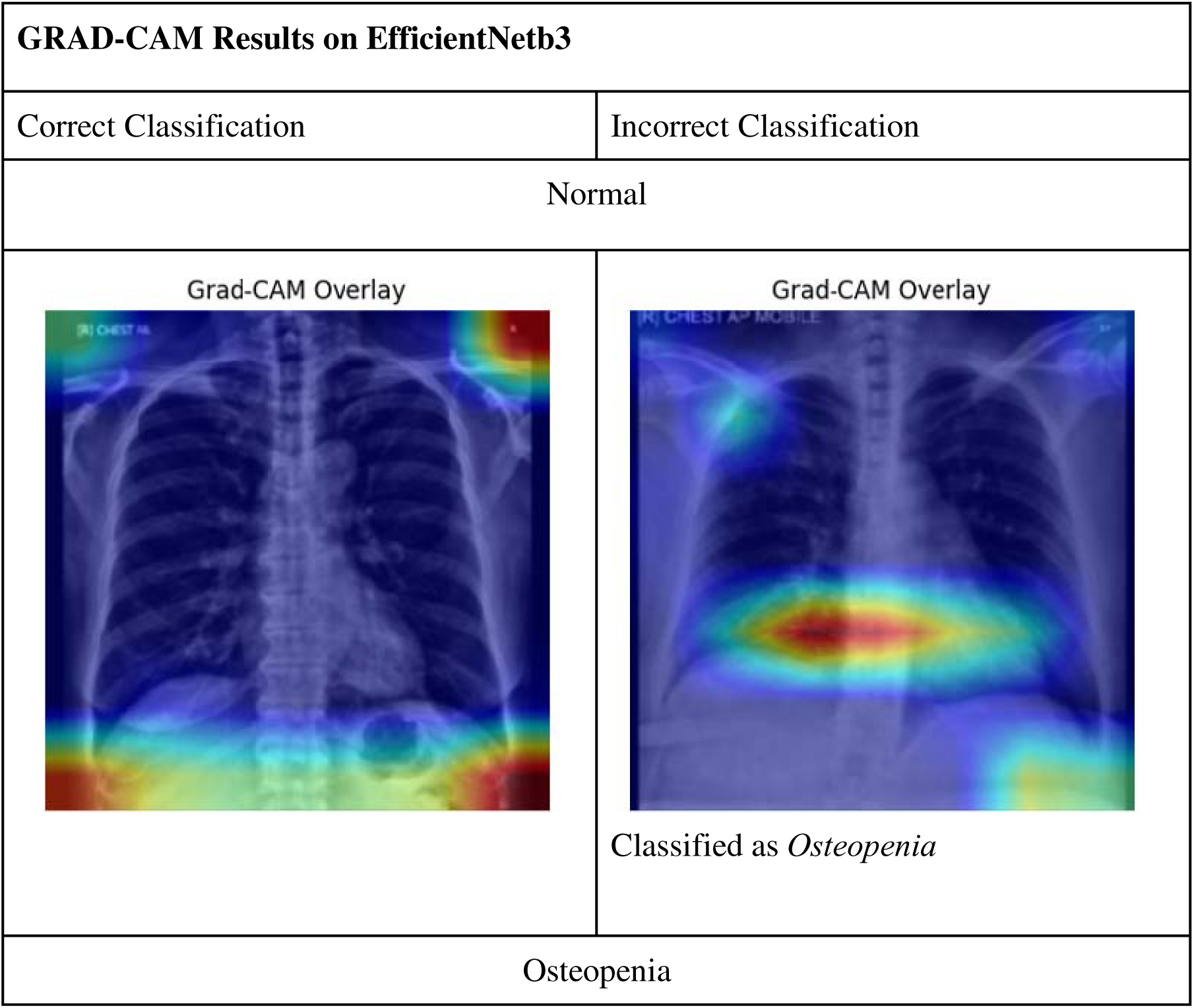
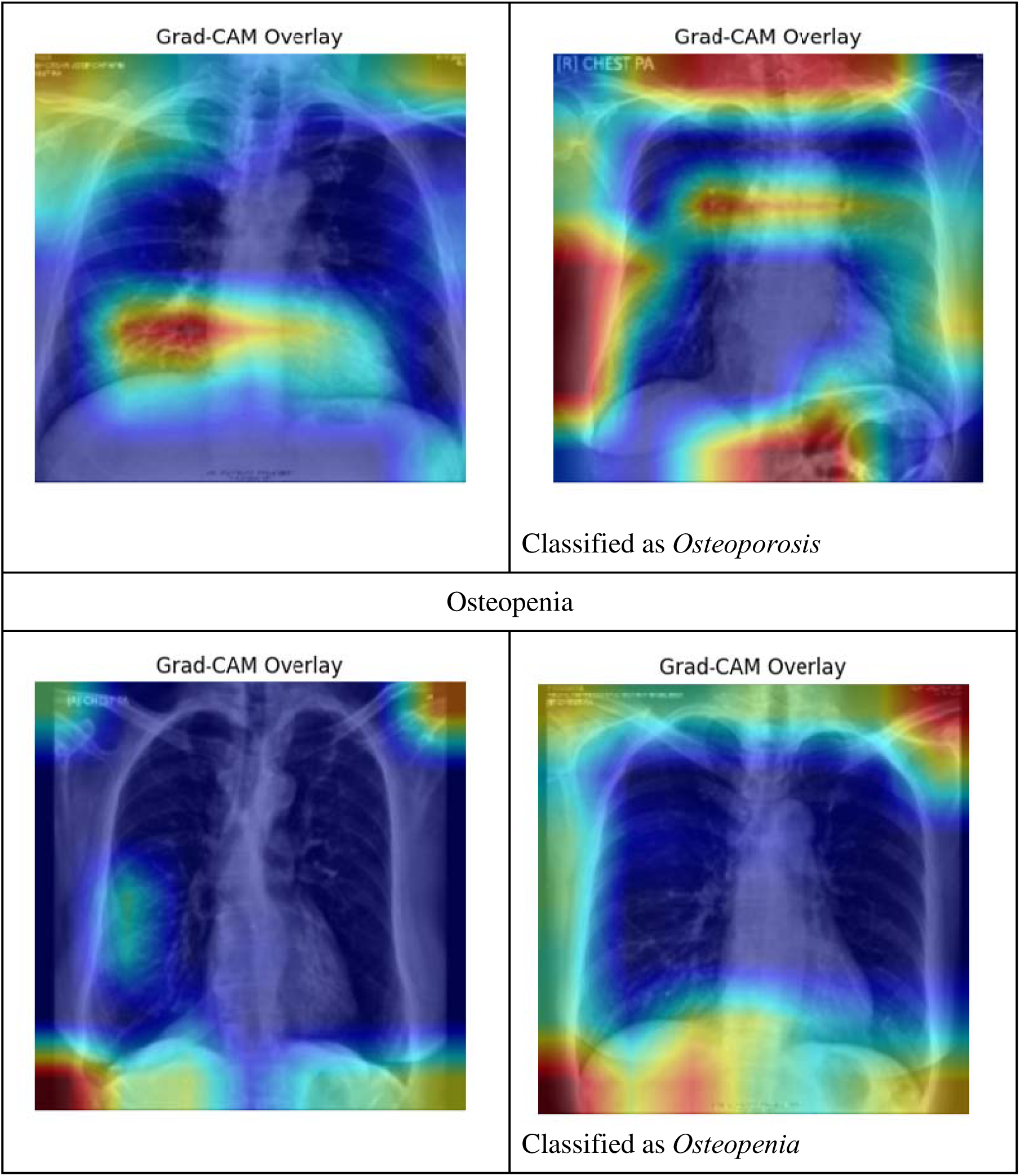
Comparison of Grad-CAM Results on the EfficientNetB3 Model Across Correct and Incorrect Classifications.

In correctly classified Normal cases, EfficientNetB3 highlights the diaphragm, posterior ribs, and upper spine, suggesting it may be learning to associate structural integrity and density patterns with normal bone health. Interestingly, in correctly classified Osteopenia cases, the model directs its attention more sharply on the central thoracic vertebrae and scapular regions— areas typically affected by early bone density loss.

For incorrectly classified cases, the attention maps are less erratic than those seen in ResNet50. Although there is some focus drift (e.g., toward the lower lungs or soft tissue margins), EfficientNetB3 still shows partial attention to relevant skeletal landmarks. This more stable attention behavior, even during misclassification, suggests better spatial prioritization and possibly stronger feature learning under multimodal constraints.

Taken together, these Grad-CAM results reinforce EfficientNetB3’s superior localization capacity and feature extraction stability, particularly when compared to ResNet50. The sharper, more clinically-aligned attention maps suggest that EfficientNetB3 can better identify disease-relevant features, potentially contributing to its improved AUC performance and reliability across multimodal inputs.

## 5 SUMMARY, CONCLUSION AND RECOMMENDATION

This study aimed to develop a multimodal deep learning-based model for osteoporosis screening by integrating clinical variables, namely, as age, and gender with chest X-ray images. The dataset comprised of chest x-ray radiographs paired with their respective clinical variables and DXA scan results of patients aged 50 and above. This study employed two convolutional neural network architectures, ResNet50 and EfficientNetb3, and compared two multimodal fusion approaches – early and late fusion.

Image preprocessing played a crucial role in improving model performance. Specifically, the application of Contrast-Limited Adaptive Histogram (CLAHE) followed by Gamma Correction enhanced bone feature visibility– specifically improving the representation of trabecular and cortical bone structures, providing the model with better cues for classification.

The image data were paired with normalized and encoded clinical variables, forming a combined feature vector used in either early-pixel level fusion or late feature-level fusion. Early fusion modified the CNN input to include the clinical features at the pixel level, while late fusion used a custom Residual MLP Classifier to interpret concatenated image and clinical features after convolution.

Model training incorporated the use of transfer learning from ChestXNet-pretrained weights and hyperparameter optimization using Optuna. The models were evaluated using Accuracy, AUC, Precision, Recall, Sensitivity, and F1 Score.

For the Global ROI, EfficientNetB3 with Late Fusion emerged as the best-performing model, yielding the highest AUC and F1 Score among all configurations. This model demonstrated the most balanced classification, especially in correctly identifying osteopenic and osteoporotic cases.

For the Local ROIs, model performance varied across anatomical regions. EfficientNetB3 consistently outperformed ResNet50 across all local ROIs—Right Clavicle, Right Scapula, and Spine. Notably, the Spine region yielded the best classification performance among the three, with Late Fusion again outperforming Early Fusion strategies. These findings highlight the value of leveraging EfficientNetB3’s compound-scaling capabilities and the discriminative power of the custom MLP used in Late Fusion.

Grad-CAM was used to visualize model attention, highlighting the anatomical areas most influential in the model’s predictions.

Despite the detailed results, several recommendations are proposed to further enhance the study’s findings. Future research should expand the dataset by incorporating larger and more diverse samples to improve model generalizability. Including additional clinical variables—such as family history, menopausal status, and history of fractures—may also enhance the model’s predictive power. Lastly, exploring hybrid fusion strategies, such as combining early and late fusion approaches or implementing voting mechanisms based on Local ROI results, could further improve classification performance.

In clinical settings, the proposed multimodal model may be integrated into radiology workflows as a decision support tool for osteoporosis risk stratification. For patients undergoing chest X-rays for unrelated conditions, the model could provide opportunistic screening by flagging high-risk cases based on both imaging and available clinical data. This allows clinicians to identify patients who may benefit from further diagnostic evaluation (e.g., DXA scans) and early intervention—especially in resource-limited environments where DXA is not routinely available.

This study provides a foundation for leveraging multimodal AI in low-resource settings where DXA is not readily accessible, ultimately contributing to early diagnosis and intervention in osteoporosis care.

## Data Availability

All data produced in the present study are available upon reasonable request to the authors

## Notes

### Competing Interest Statement

The authors have declared no competing interest.

### Author Declarations

Ethics IRB of The Medical City gave ethical approval for this work

### Summary of Updates

rewrote the abstract to satisfy the word number requirement.

